# *FRMPD4*, a causal gene for intellectual disability and epilepsy, is associated with X-linked non-syndromic hearing loss

**DOI:** 10.64898/2026.03.27.26349271

**Authors:** Daniel Liedtke, Kristen Rak, Katrina M Schrode, Philip Hehlert, Niloofar Chamanrou, Daniel Bengl, Radoslaw Katana, Soganad Heydaran, Julia Doll, Mei Han, Indrajit Nanda, Pingkalai R Senthilan, Lukas Jürgens, Linda Bieniussa, Johannes Voelker, Cordula Neuner, Michaela AH Hofrichter, Jörg Schröder, Renske T.W. Schellens, Erik de Vrieze, Erwin van Wijk, Ulrich Zechner, Stefan Herms, Per Hoffmann, Tobias Müller, Marcus Dittrich, Oliver Bartsch, Peter M Krawitz, Eva Klopocki, Wafaa Shehata-Dieler, Reza Maroofian, Tao Wang, Paul F Worley, Martin C Göpfert, Hamid Galehdari, Amanda M Lauer, Thomas Haaf, Barbara Vona

## Abstract

**Background:** Understanding the phenotypic spectrum of disease-associated genes is essential for accurate diagnosis and targeted therapy. *FRMPD4* (FERM and PDZ Domain Containing 4) has previously been associated with intellectual disability and epilepsy. However, its potential role in non-syndromic hearing loss has not been explored.

**Methods:** We performed genetic analysis in two unrelated families presenting with non-syndromic sensorineural hearing loss, identifying maternally inherited missense variants in *FRMPD4*. Clinical phenotyping included audiological assessment and evaluation for neurodevelopmental involvement. Cross-species expression analyses were conducted in *Drosophila*, zebrafish, and mouse. Functional characterization included quantitative evaluation of sound-evoked responses in *Drosophila nicht gut hörend* (*ngh*) mutants, assessment of neuronal development and acoustic startle responses in zebrafish loss of function models, and morphological cochlear analyses with auditory brainstem response measurements in knockout mice.

**Results:** Three affected males from two unrelated families presented with prelingual, bilaterally symmetrical sensorineural hearing loss, with confirmed congenital onset in one individual and no evidence of neurodevelopmental abnormalities. Cross-species analyses demonstrated evolutionarily conserved expression of *FRMPD4* in auditory structures. In *Drosophila*, quantitative analysis of sound-evoked responses in *ngh* mutants revealed impaired auditory function. Zebrafish loss of function models exhibited reduced neuronal populations in the otic vesicle and posterior lateral line, abnormal neuromast development, and diminished acoustic startle responses. In mice, *Frmpd4* knockout resulted in high-frequency hearing loss and cochlear abnormalities consistent with the human phenotype.

**Conclusions:** Our findings expand the phenotypic spectrum of *FRMPD4* to include non-syndromic sensorineural hearing loss and establish its evolutionarily conserved role in auditory function. These results have direct implications for genetic diagnosis and variant interpretation in patients with hearing loss.

## Background

Hereditary non-syndromic hearing loss is one of the most common and genetically heterogeneous sensory disorders. It affects approximately one to two per 1000 newborns, with genetic factors accounting for the majority of cases (Morton and Nance 2006). Autosomal recessive and dominant forms together account for roughly 95% of inherited non-syndromic hearing loss (Vona et al. 2020). Of the more than 155 genes currently associated with non-syndromic hearing loss, only five map to the X chromosome (*AIFM1, POU3F4*, *COL4A6*, *PRPS1*, and *SMPX*). Accordingly, X-linked forms are very rare and are estimated to account for 2-5% of non-syndromic hearing loss (Vona et al. 2020).

Genetic studies of non-syndromic hearing loss have uncovered numerous examples of pleiotropy, in which genes initially implicated in syndromic or neurodevelopmental disorders also contribute to isolated auditory phenotypes (Murray et al. 2017; Rehman et al. 2014; Riazuddin et al. 2009; Richard et al. 2021). These findings highlight shared molecular pathways between the auditory system and other organ systems and underscore the importance of reevaluating the phenotypic spectrum of disease-associated genes as new functions which are revealed through comprehensive genetic and functional studies (Tshering et al. 2025; Vona 2024).

The evolutionary conservation of many hearing-associated genes across vertebrate species further supports their essential role in auditory function. These genes frequently exhibit conserved expression in auditory organs, and functional studies have shown that their encoded proteins are essential for species-specific auditory function (Elkon et al. 2015; Pei et al. 2016). This suggests a common evolutionary origin for key auditory mechanisms and reveals conserved cellular functions (Carey and Amin 2006; Fritzsch and Straka 2014; Streit 2001). Beyond rodent models, other organisms such as zebrafish (*Danio rerio*) (Haddon and Lewis 1996; Raible and Kruse 2000) and *Drosophila melanogaster* (Li et al. 2018) have become powerful systems for studying gene function in hearing, despite species-specific adaptations in hearing organ structure and neuronal sound processing. For example, sound detection in zebrafish partly relies on neuromast hair cells of the anterior and posterior lateral line, which sense fluid movement along the body axis. These hair cells are homologs of the inner-ear hair cells and are evolutionarily related not only to hair cells in the human organ of Corti but also to the chordotonal mechanosensory neurons mediating hearing in *Drosophila* (Fritzsch and Straka 2014; Hassan and Bellen 2000; Nicolson 2017; Nicolson 2005).

In this study, we identify two novel missense variants in *FRMPD4* (OMIM: 300838) through exome sequencing and analysis of two previously undiagnosed families with non-syndromic hearing loss. *FRMPD4* has been linked to schizophrenia and intellectual disability (Hu et al. 2016; Matosin et al. 2016; Trujillano et al. 2017) and plays a functional role in dendritic outgrowth and morphogenesis (Lee et al. 2008). It has also been shown to interact with whirlin as part of the Usher syndrome type 2 protein complex in photoreceptor cells (Schellens et al. 2022), and *FRMPD4* pathogenic variants have been associated with isolated epilepsy and epilepsy with intellectual disability (Li et al. 2024). Here, we demonstrate that FRMPD4 additionally plays a conserved role in auditory pathways across humans, *Drosophila*, zebrafish, and mice, expanding its functional repertoire and implicating it in non-syndromic hearing loss.

## Methods

### Patient recruitment and clinical assessment

Families were recruited through a large rare disease study and through data sharing with collaborating clinicians. The sole inclusion criterion was the presence of hereditary hearing impairment. This study was approved by the Medical Faculty at the University of Würzburg (approval number: 46/15). Written informed consent was obtained from all participating individuals or their parental guardians prior to enrollment.

Otolaryngologic, audiological, and general medical data were ascertained from the medical records of both families and are described in detail in the Supplemental Materials and Methods.

### Molecular genetic work-up, exome sequencing, and data analysis

Genomic DNA (gDNA) was extracted from whole blood of affected individuals and available family members using a standard salt extraction method (Family 1: II:2, II:3, III:2, and III:3; Family 2: III:1, III:2, III:3, and IV:1). For Family 1, genome-wide SNP genotyping was performed using the Illumina Omni1-Quad SNP-array (Illumina, San Diego, CA, USA) according to the manufacturer’s specifications and analyzed as previously described (Vona et al. 2014). Subsequently, the gDNA of both affected children from Family 1 underwent targeted sequencing of an 80-gene deafness panel including genes associated with both syndromic and non-syndromic hearing loss. Sequencing was performed on an Illumina HiSeq2000 platform by Otogenetics Corporation (Norcross, GA, USA) and data were analyzed as described previously (Vona et al. 2014). Exome sequencing was performed on gDNA from the two affected boys and their parents from Family 1 (II:2, II:3, III:2, and III:3) using the SeqCap EZ Human Exome Library v3 (64M) enrichment kit (Roche NimbleGen). Libraries were sequenced as 2 × 100 bp paired-end reads on an Illumina HiSeq 2000 platform (Life and Brain GmbH, Bonn, Germany). For family 2, an exome library from the proband (IV:1) was prepared using the Agilent SureSelect v6 kit (Agilent, Santa Clara, CA, USA) and exome sequenced on an Illumina HiSeq4000 platform.

Exome data from Family 1 were aligned to the human reference genome (GRCh37/hg19) using the Cologne Center for Genomics Varbank v2.1 pipeline. This pipeline incorporates GATK for base recalibration, local alignment, and variant score recalibration. Variant calling was performed using MPILEUP, GATK and DINDEL according to best practice recommendations (McKenna et al. 2010). GeneTalk was additionally used for variant filtering and result validation (Kamphans and Krawitz 2012). Exome data from individuals in Family 2 was demultiplexed and aligned to the human reference genome (GRCh38) using Burrows-Wheeler Aligner for subsequent variant calling. Detailed exome filtering strategies, analysis workflows, and variant prioritization are described in the Supplemental Materials and Methods.

### Animal maintenance and experimentation

#### Drosophila

Fly stocks were maintained at 25°C on a standard cornmeal-agar diet. The mutants were obtained from the Bloomington Stock Center (BL 60756; genotype: *y^1^ w*; Mi{MIC}CG42788^MI02203^*). Reporter knockout (KO) line CG42788^1,Gal4,3xP3>DsRed^ (generated in this study) was used to investigate expression in Johnston’s organ in the second antennal segment.

Methods related to RT-PCR, immunofluorescence staining, and electrophysiological and mechanical recordings in *Drosophila* are included in the Supplemental Materials and Methods.

### Zebrafish

Zebrafish (*Danio rerio*) were bred and maintained in the aquatic facilities of the Biocenter of the Julius-Maximilians-University Würzburg, Germany according to FELASA guidelines (Aleström et al. 2020) (husbandry permit number 568/300-1870/13). Adult fish were kept at a mean temperature of 24-26°C in 10 l glass and 2.5 l plastic tanks, while embryos younger than 120 h post-fertilization (hpf) were raised at a temperature of 28.5°C in an incubator. A daily light cycle of 10 h dark/14 h light was maintained for breeding fish. Preconditioned reverse osmosis water with adjusted conductivity 500-1,100 µS/cm, pH 7.0 and stable water hardness was used. A food combination of *Artemia nauplii* and GEMMA Micro Food (age dependent sizes; Skretting, USA) was standard. All experimental procedures were performed according to the guidelines of the German animal welfare law and approved by the local government (Government of Lower Franconia; Tierschutzgesetz §11, Abs. 1, Nr. 1; Genotyping and startle response protocol permit number: DMS-2532-2-9 and DMS-2532-2-428). Zebrafish embryos (*Danio rerio*) of the *AB/TU* (ZDB-GENO-010924-10) and *AB/AB* (ZDB-GENO-960809-7) strains were used in this study and were staged by morphological characteristics (Kimmel et al. 1995). *frmpd4^sa12377^*mutants were obtained from the European Zebrafish Resource Center (KIT, Karlsruhe, Germany; allele name: sa12377; ZFIN line ID: ZDB-ALT-130411-2117) and possess a G>T variant at an essential splice site in exon 11.

Methods related to whole mount *in situ* hybridization, Morpholino generation, CRISPR/Cas9 gene editing, RNA rescue via overexpression, DASPEI staining, immunofluorescence, scanning electron microscopy, and startle response testing with analysis are included in the Supplemental Materials and Methods.

### Mouse

Mouse breeding and procedures were conducted in strict accordance with NIH Guide for Care and Use of Laboratory Animals. Mouse studies were carried out in accordance with the Guide for the Care and Use of Laboratory Animals, the ARRIVE guidelines (Percie du Sert et al. 2020) and Johns Hopkins University Animal Care and Use Committee. Conditional KO *Frmpd4^-/-^* mice (this mouse model was originally called *Preso1*^-/-^, hereafter called *Frmpd4^-/-^* for simplicity) were generated as previously described and were viable, fertile, and showed similar development and breeding behaviors as wild type (WT) littermates (Hu et al. 2012). Briefly, exon 3 was deleted by inserting flanking *loxP* sites and a *loxP*/PGK-neo cassette and crossing with *CMV-cre* mice. The transgenic colony was maintained on a C57BL/6J background. Genotyping was performed as previously described using PCR of tail DNA (Hu et al. 2012). Mice were housed in temperature-controlled rooms with 12 h light/dark cycle and had free access to food and water.

Mouse experimental methods related expression analysis of *Frmpd4* in the cochlea and other tissues, auditory brainstem response (ABR) testing and analysis are described in Supplemental Materials and Methods. Primer sequences are listed in Supplementary Table 1.

### Statistical analysis

Relative distances of posterior lateral line primordium (PLL) migration, neuromast numbers in the cranial lateral line, and startle response reaction times were analyzed using OriginPro 2021 (OriginLab Corporation, Northampton, MA, USA) and visualized using boxplot/data point diagrams. Diagrams show whiskers indicating standard deviation (coefficient value: 1.5), the median (parallel line), the mean value (small box), and the upper and lower quartile (large box). For statistical analysis, the obtained data values were first tested for normal distribution (ANOVA test), and the significance was determined using a two-tailed Mann-Whitney-U test. An asterisk indicates significant changes between groups U < 0.01, while n.s. marks not significantly different groups. Generalized linear mixed model analysis was performed using the nlme package in R (Pinheiro et al. 2022; Vazquez et al. 2010). The model included frequency and genotype as factors and their interaction, as well as a random effect to account for repeated measures. Post hoc Tukey tests were computed to evaluate differences between genotypes by frequency. A *p*-value < 0.05 was considered statistically significant.

## Results

### Clinical evaluation of Family 1

Family 1 consists of a three-generation non-consanguineous family from Europe (Figure 1A). The youngest generation comprises three children, two of whom are affected by hearing loss. The youngest child (III:3) was born after an uneventful pregnancy. Following a failed newborn hearing screening, the child was diagnosed with mild-to-moderate sensorineural hearing loss shortly thereafter. The hearing of his older siblings was retrospectively evaluated, identifying mild sensorineural hearing loss in III:2. The older siblings were born after a complicated pregnancy after 35 weeks of gestation. Each had a birth weight of 2,120 g (19th percentile, Z-score: -0.88) and a body length of 46 cm (34th percentile, Z-score: -0.40). Individual III:1 was diagnosed with delayed speech and language development and received speech therapy. Otoscopy performed at the age of 6-10 years revealed normal closed tympanic membranes. Pure-tone audiometry and bilateral otoacoustic emissions were normal and reproducible. The most recent hearing test at 6-10 years of age confirmed normal hearing (data not shown), further arguing against a mitochondrial mode of inheritance.

**Figure 1.**
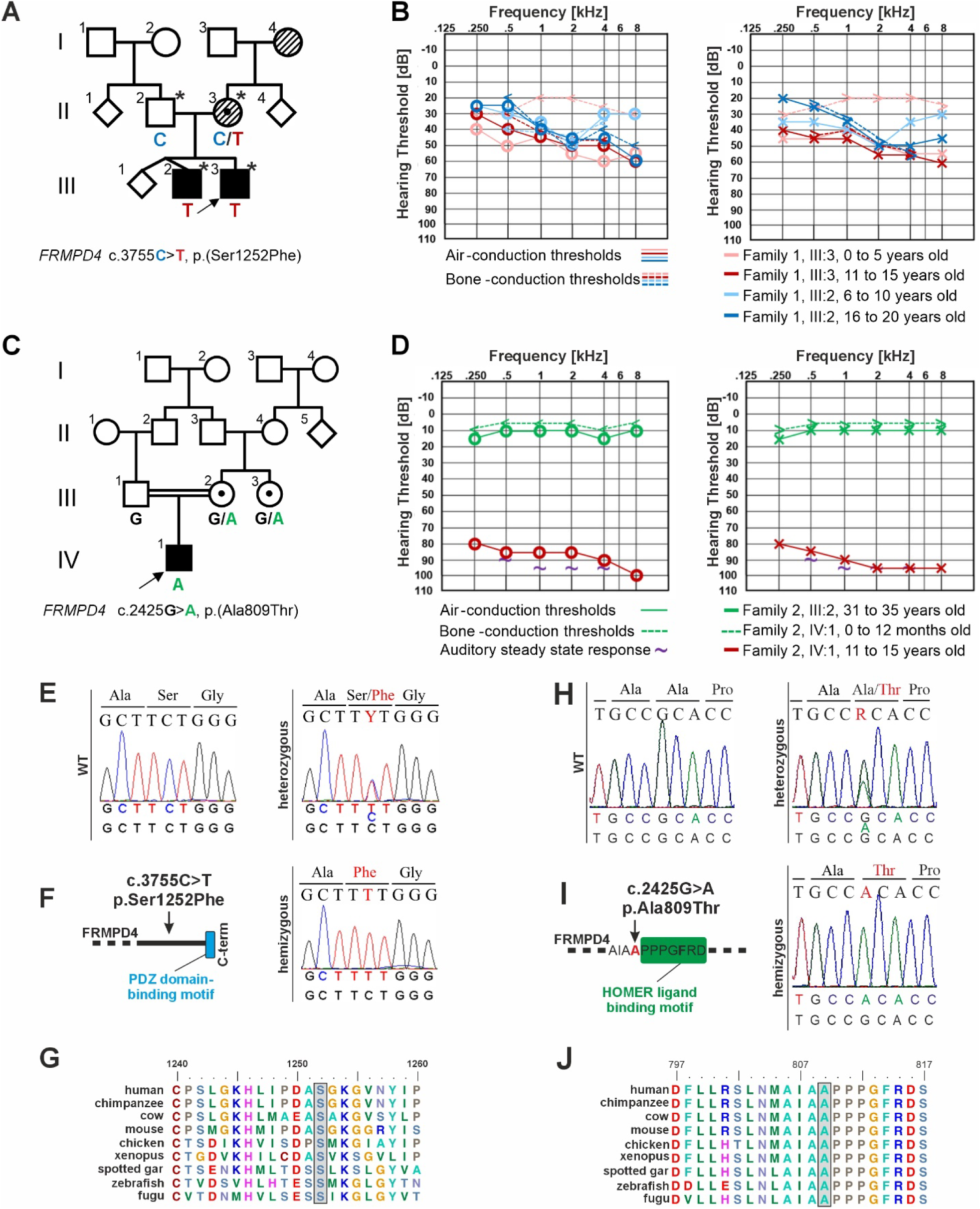
Identification of the putative disease-causing *FRMPD4* variant in two families with non-syndromic hearing loss. **(A)** Pedigree of family 1. Black symbols are affected individuals, unfilled symbols are unaffected individuals and individuals with stripes have late-onset hearing loss. Asterisks denote the family members whose exomes were sequenced. Genotypes at the c.3755C>T, p.(Ser1252Phe) position are denoted below each pedigree symbol. **(B)** Pure-tone audiograms of the affected males. Unmasked bone conduction measurements are included when available. Audiograms from III:3 at the age of 0-5 years (pink) and 11-15 years (red). Audiograms from III:2 at age 6-10 years (light blue) and 16-20 years (dark blue). Air- and bone-conduction thresholds are represented with circles and <, as well as crosses and > for right and left ears, respectively. **(C)** Pedigree of family 2 with genotypes of the *FRMPD4* c.2425G>A, p.(Ala809Thr) variant below each individual in whom segregation analysis was performed. **(D)** Audiometry from the proband (IV:1) included auditory steady-state responses at the age of 0-12 months (purple tilde), as well as a most recent pure-tone audiogram at the age of 11-15 years is in red. Pure-tone audiometry of the mother of the proband (III:2) at the age of 31-35 years is in green. **(E)** Direct sequencing confirmed the *FRMPD4* c.3755C>T, p.(Ser1252Phe) variant in family 1. The unaffected father was wild type (WT), the mother with self-reported mild hearing loss was heterozygous, and the two affected children were hemizygous. **(F)** The c.3755C>T, p.(Ser1252Phe) variant impacts an amino acid residue before the PDZ domain-binding motif. **(G)** Amino acid alignment of the p.Ser1252 region shows high evolutionary conservation in vertebrates. **(H)** Direct sequencing confirmed the *FRMPD4* c.2425G>A, p.(Ala809Thr) variant in family 2 with the mother of the proband (III:2) and maternal aunt (III:3) as heterozygous, the father of the proband (III:1) showing the WT allele, and the male proband with a hemizygous allele for the *FRMPD4* c.2425G>A, p.(Ala809Thr) variant. **(I)** The c.2425G>A, p.(Ala809Thr) variant impacts an amino acid residue directly before the HOMER ligand binding motif. **(J)** Amino acid alignment of the p.Ala809 region shows high evolutionary conservation in vertebrates.

Annual or semi-annual otoscopy and audiometric follow-up have been performed in both affected children. Bilateral distortion product and transient evoked otoacoustic emissions showed no reproducible responses in these individuals. Individual III:2 required early childhood speech therapy related to a delayed diagnosis. At most recent evaluation, hearing loss was moderate in individual III:3 (PTA_0.5-4kHz_ [pure-tone average] 46.25 dB HL [hearing level] [right] and PTA_0.5-4kHz_ 50 dB HL [left]), and mild in individual III:2 (PTA_0.5-4kHz_ 38.75 dB HL [right] and PTA_0.5-4kHz_ 40 dB HL [left]) at the age of 10-20 years, respectively (Figure 1B). Hearing loss was bilaterally symmetrical and stable over time. Speech audiometry without hearing aids performed in III:3 disclosed a 30% and 40% discrimination for left and right ears, respectively, that improved to 80% and 90% with hearing aids. Free-field audiometry without hearing aids revealed responses at stimulus levels between 60 and 70 dB. Hearing aids effectively rehabilitated hearing loss in both individuals. There has been no dizziness, ear pain, or ear discharge. Both attended mainstream schools, demonstrated good academic performance, and showed no concentration difficulties. Their school histories do not support developmental delay and intellectual disability. Pediatric developmental milestones followed an age-appropriate course. Whole blood analytics and electrocardiogram results were unremarkable. Clinical examination confirmed the absence of additional phenotypic abnormalities.

The mother (II:3) reported noticing mild hearing loss in her fourth decade of life, and the maternal grandmother (I:4) required hearing aids at age 55-60 years. Audiograms from both individuals are unavailable.

### Clinical evaluation of Family 2

Family 2 consists of a four-generation pedigree of Middle Eastern (Figure 1C). The proband (IV:1) was reported to have congenital bilateral non-syndromic hearing loss. Auditory steady-state response testing at 0-12 months of age estimated hearing thresholds of 90 dB bilaterally with an 85% confidence level (Figure 1D). Stimuli consisted of 100% amplitude-modulation and 20% frequency-modulated tones between 80 to 100 dB HL in 5 dB interval steps using headphones. Thresholds were estimated at 0.5, 1, 2, and 4 kHz in both ears. Otoacoustic emissions were absent. The child received cochlear implants in the prelingual stage. Pure tone audiometry at the age of 11-15 years confirmed severe sensorineural hearing loss (PTA_0.5-4kHz_ 86.25 dB HL) (Figure 1D). Tympanometric evaluation showed normal middle ear function. The proband’s mother (III:2) was confirmed to have normal hearing at the age of 30-35 years (Figure 1D), with speech recognition thresholds at 10 dB HL bilaterally and speech discrimination scores of 96% (right) and 92% (left).

### Exome sequencing identifies *FRMPD4* variants in patients with non-syndromic hearing loss

**Family 1:** Genetic investigation was initiated following routine molecular diagnostic evaluation that failed to yield an informative diagnosis. This included analysis of *GJB2* and *STRC* with multiplex ligation-dependent probe amplification diagnostic testing of *GJB2*, *GJB3*, *GJB6*, *POU3F4* and *WFS1* (MRC-Holland). SNP-array analysis of the two affected individuals yielded an uninformative result (Illumina Omni1-Quad). An 80-gene deafness panel was subsequently performed but did not establish a genetic diagnosis.

gDNA from the parents and two affected children (II:2, II:3, III:2, and III:3) was subjected to exome sequencing (Figure 1A). Variant filtering disclosed 304,240 variants that were systematically filtered under autosomal recessive, autosomal dominant, and X-linked inheritance patterns (Supplementary Table 2). Fourteen heterozygous autosomal variants inherited from an unaffected parent were excluded (Supplementary Table 3). Analysis of variants in hearing loss-associated genes uncovered a heterozygous variant in *USH1C* (NM_153676.3:c.1591C>T, p.Arg531Cys), a gene causal for autosomal recessive Usher syndrome type 1 with congenital hearing impairment. This variant was classified as a variant of uncertain significance (VUS; PM2_P) and was excluded due to absence of a second variant and lack of clinical concordance (Supplementary Table 4). A likely benign (PM2_P, BP4_S) heterozygous *BDP1* (NM_018429.2:c.2351A>G, p.Lys784Arg) variant was also identified. Further exome-wide analysis uncovered a likely benign (PP3_P, BS2_S) hemizygous variant in *PHKA2* (NM_000292.2:c.202G>A, p.Asp68Asn) that was transmitted from the mother to both affected children. A heterozygous VUS (PM2_P, PP3_P) in *MYBPH* (NM_004997.2:c.989T>G, p.Leu330Arg) was identified. This gene has so far been characterized as a modifier of hypertrophic cardiomyopathy (Mouton et al. 2016) and with a putative skeletal muscle function (Mead et al. 2024) but no known association with hearing loss. A single missense variant in *FRMPD4* NM_001368397.1:c.3755C>T (p.Ser1252Phe) was identified in a hemizygous state in the two affected boys and was heterozygous in their mother that was confirmed with Sanger sequencing (Figure 1E). This variant is absent in gnomAD (n = 730,947 exomes and 76,215 genomes), TopMed (n = 138,000 genomes), the All of Us (n = 414,840 genomes), and the Varbank in house exome database (n = 511 exomes). *In silico* variant effect predictors suggested a nearly unanimous deleterious effect (Table 1). The cytosine at position c.3755 is highly conserved (phyloP: 4.64 [-14.1 to 6.4]) and occurs before the PDZ domain binding motif (Figure 1F). Similarly, the serine-to-phenylalanine substitution alters a highly conserved amino acid residue (Grantham distance: 155 [0-215]), with conservation observed in 48 of 75 species examined, including zebrafish (*Danio rerio*) and mouse (Figure 1G).

**Table 1.**
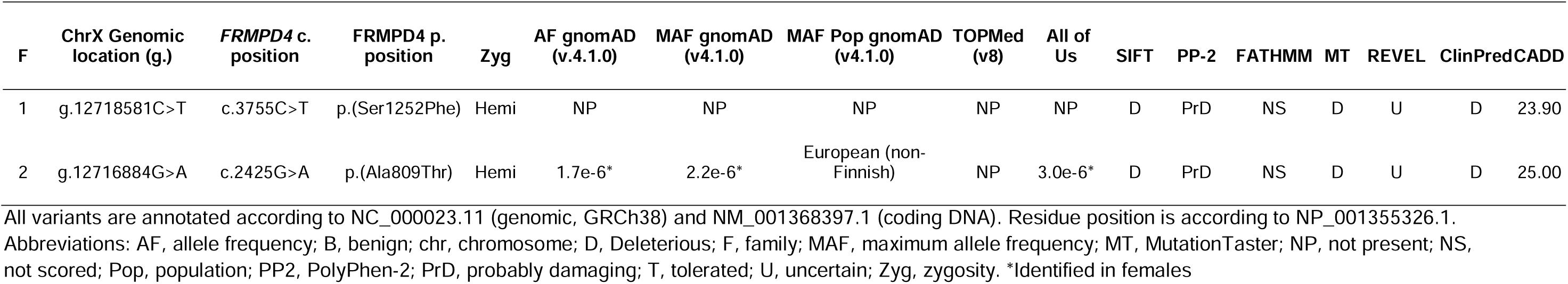
FRMPD4 variants identified in hearing impaired individuals.

| F | ChrX Genomic location (g.) | *FRMPD4* c. position | *FRMPD4* p. position | Zyg | AF gnomAD (v.4.1.0) | MAF gnomAD (v4.1.0) | MAF Pop gnomAD (v4.1.0) | TOPMed (v8) | All of Us | SIFT | PP-2 | FATHMM | MT | REVEL | ClinPred | CADD |
| --- | --- | --- | --- | --- | --- | --- | --- | --- | --- | --- | --- | --- | --- | --- | --- | --- |
| 1 | g.12718581C>T | c.3755C>T | p.(Ser1252Phe) | Hemi | NP | NP | NP | NP | NP | D | PrD | NS | D | U | D | 23.90 |
| 2 | g.12716884G>A | c.2425G>A | p.(Ala809Thr) | Hemi | 1.7e-6* | 2.2e-6* | European (non-Finnish) | NP | 3.0e-6* | D | PrD | NS | D | U | D | 25.00 |

All variants are annotated according to NC\_000023.11 (genomic, GRCh38) and NM\_001368397.1 (coding DNA). Residue position is according to NP\_001355326.1.
Abbreviations: AF, allele frequency; B, benign; chr, chromosome; D, Deleterious; F, family; MAF, maximum allele frequency; MT, MutationTaster; NP, not present; NS,
not scored; Pop, population; PP2, PolyPhen-2; PrD, probably damaging; T, tolerated; U, uncertain; Zyg, zygosity. \*Identified in females

**Family 2:** Exome sequencing of the proband of Family 2 (IV:1) uncovered 98,819 variants that were filtered according to all possible inheritance patterns (Supplementary Table 5). Analysis of hearing loss-associated genes uncovered a heterozygous *CDH23* (NM_022124.5:c.9014C>T, p.Ala3005Val) variant that was classified as VUS (PM2_P). Exome-wide analysis identified three homozygous variants in *SYPL2* (NM_001040709.1:c.406_407dup, p.Leu137Thrfs*21), *MAPKAPK5* (NM_139078.2:c.1099G>A, p.Gly367Ser), and *TNRC6C*

(NM_001142640.2:c.2483G>T, p.Gly828Val) in genes that are not expressed in the inner ear, or associated with other phenotypes (Supplementary Table 6). Two further homozygous variants were identified in uncharacterized genes that were excluded from consideration on the basis of weak variant effect prediction scores (Supplementary Table 6). A single variant in *FRMPD4* (NM_014728.3:c.2425G>A, p.Ala809Thr) emerged as a candidate variant (Table 1, Supplementary Table 4). Presence of two independent families with missense variants in *FRMPD4* prompted further expression and functional analysis to associate *FRMPD4* with non-syndromic hearing loss.

### Comparison of the *FRMPD4* variants associated with non-syndromic hearing loss with variants causing intellectual disability

*FRMPD4* has a Residual Variation Intolerance Score of -1.84 ranking *FRMPD4* in the top 2% of genes most intolerant to variation. So far, 13 variants in *FRMPD4* have been associated with intellectual disability of varying severity, isolated epilepsy, or epilepsy with intellectual disability, the vast majority of which are maternally inherited (Supplementary Table 7), and so far, there is no clear distinct genotype-phenotype association. The two novel variants we describe that are associated with non-syndromic hearing loss do not appear to cluster to any one region of the gene or protein domain. Similarly, the previously described variants also appear to impact positions along the entire protein. A schematic overview of variant localization in the FRMPD4 protein is presented in Supplementary Figure 1.

**> Supplementary Figure 1**

### FRMPD4 orthologues are expressed in hearing organs during development

The high conservation of FRMPD4 variants identified in patients implied a potential function on hearing organs in other vertebrates. Genomic comparison between FRMPD4 orthologues showed relatively high amino acid conservation in the two functional domains, the PDZ and the FERM domain (Supplementary Figures 2A and 3). While mice share up to 99.3% amino acid-identity to the human protein domains, the zebrafish (up to 89.7% amino acid-identity) and even the *Drosophila* (up to 58.9% amino acid-identity) orthologues show remarkable conservation values. Synteny analyses further indicated the prolonged evolutionary conservation of the *frmpd4* locus in the investigated species (Supplementary Figure 2B).

**> Supplementary Figures 2 and 3**

To further test auditory-specific expression of FRMPD4 orthologues in vertebrates we investigated different animals for the presence of transcripts in their corresponding organs. *Frmpd4* transcripts in the mouse could be detected via qPCR in several neuronal tissues and kidney, but also in auditory organs, such as the cochlea and the cochlear nucleus (Figure 2A). *Frmpd4* was expressed nearly exclusively in the spiral ganglion neurons of the mouse at postnatal day 8 (P8). Expression in the spiral ganglion neurons increases between embryonic to early postnatal stages where it remains stably expressed between P8 and P30. Spiral ganglion neuron expression appears across types Ia, Ib, Ic and type 2 spiral ganglion neurons (Supplementary Figure 4). Validation of FRMPD4 localization by immunofluorescence on mouse cochlea supported the presence of FRMPD4 in organ of Corti, in the spiral ganglion and in the acoustic nerve during development (Figure 2B; Supplementary Figure 4).

**Figure 2.**
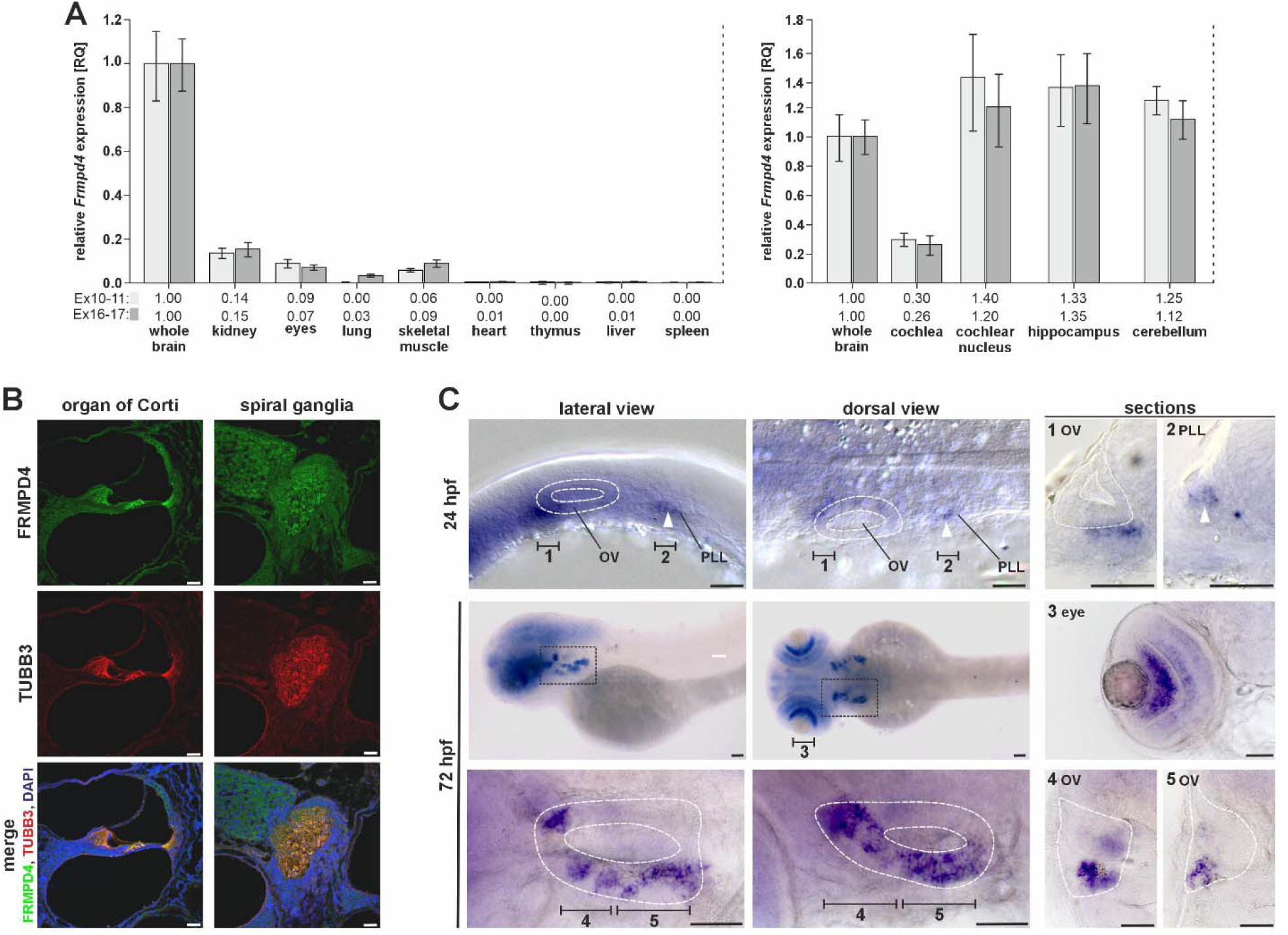
Frmpd4 expression in mice and zebrafish **(A)** Expression of *Frmpd4* in adult mice was detected mainly in whole brain, eyes, kidney, skeletal muscle, and in hearing organ tissues via qPCR. Expression in specific neuronal regions associated with hearing imply a relatively low *Frmpd4* expression in cochlear cells. **(B)** Localization of FRMPD4 and TUBB3 by immunofluorescence in cross-sections in the organ of Corti and in the spiral ganglion show very specific localization of FRMPD4 in distinct cell types, like phalangeal and hair cells (5 µm section; age: 3 mo). **(C)** Expression of *frmpd4* in zebrafish was investigated by *in situ* hybridization and was detected in the otic vesicle (OV; white dashed line) and in the posterior lateral line primordium (PLL; white arrowhead). Sectioning planes are indicated by black bars and their corresponding number (1: anterior otic vesicle 24 hpf; 2: lateral line primordium 24 hpf; 3: eye; 4: posterior otic vesicle 72 hpf; 5: anterior otic vesicle 72 hpf). Scale bars in B and C indicate 50 µm

**Figure 3.**
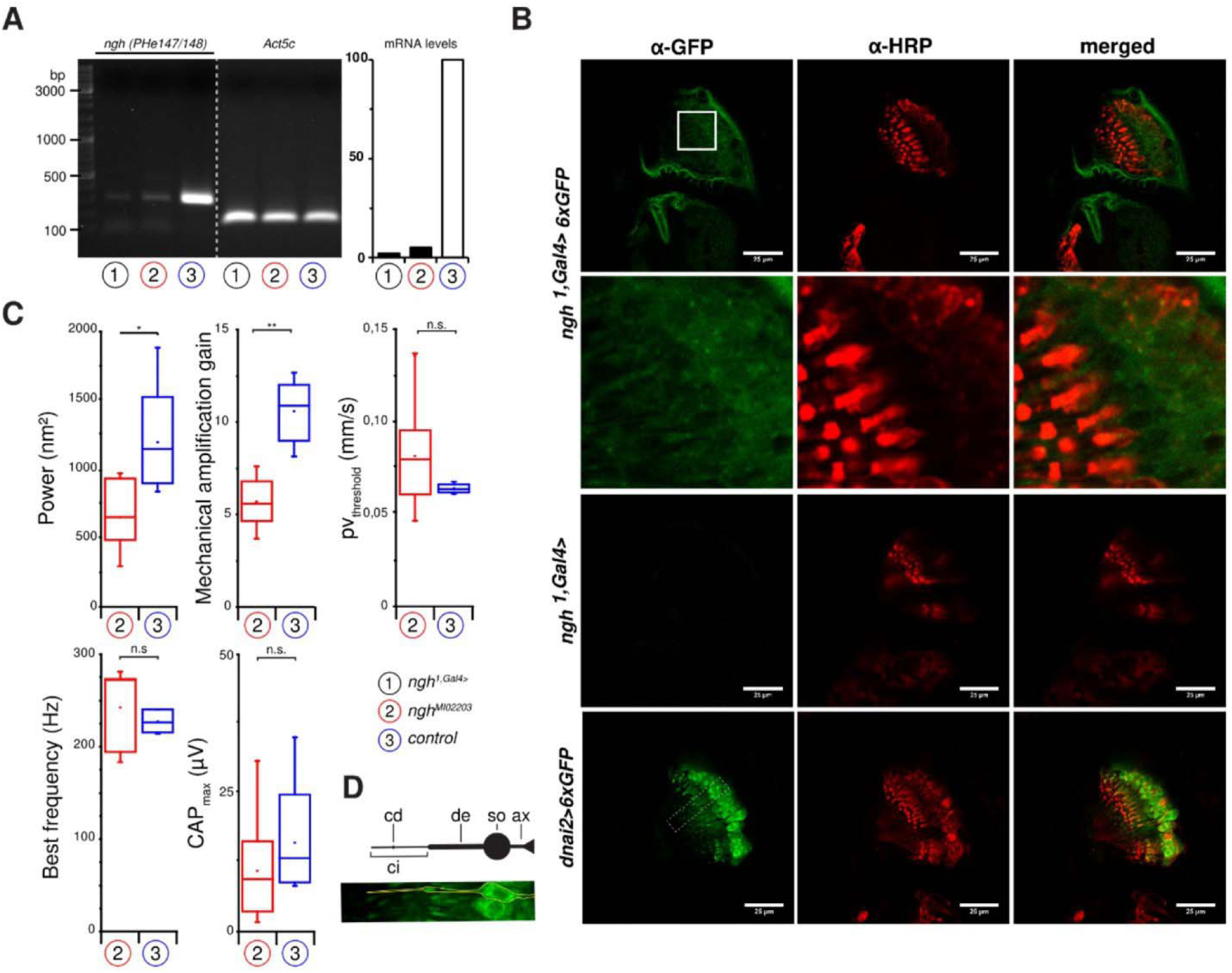
FRMPD4 function in hearing is evolutionary conserved in invertebrates. **(A)** RT-PCR analysis of *ngh* expression (*ngh^1,Gal4^*; marked in black) and ENU mutants (*ngh^MI02203^*; marked in red) in comparison to wild type controls (*w^1118^*; marked in blue). *Act5c* was used as reference. **(B)** Expression analysis of *ngh* utilizing a Promotor-trap generated via CRISPR (*ngh^1,Gal4^*) and hexameric cytoplasmic GFP (6xGFP) as reporter gene. Weak expression of *ngh* (*ngh^1,Gal4>^*6xGFP) could be detected in JONs (*dnai2>6xGFP* as positive control). Scale bars indicate 25µm. **(C)** Hearing was quantified by measuring the resonance frequency of the mechanical free fluctuations of the antennal sound receiver, the power of these free fluctuations, the nonlinear amplification gain provided by JON motility, and sound particle velocity required to evoke electrical JON compound action potentials (CAPs), and the maximum amplitude of these CAPs. Statistical significance was calculated with Mann-Whitney U tests. Asterisks indicate significant differences between genotypes (*n.s.=* not significant). **(D)** Schematic view of the morphology of mechanosensitive JO neurons: ax (axon), so (soma), de (dendrite), ci (cilium), cd (ciliary dilation).

**>Supplementary Figure 4**

Investigation of *frmpd4* expression in zebrafish was performed by whole-mount *in situ* hybridization to resolve spatio-temporal expression pattern changes during embryonic and early larval development (Figure 2C). *frmpd4* expression was detected in a broad, undefined expression domain in neuronal structures 24 hpf (hours post fertilization). Besides these domains, *frmpd4* transcripts were detected in cell clusters adjacent and anterior to the otic placode/otic vesicle (region 1 in Figure 2C) and in the lateral line primordium (region 2 in Figure 2C). Both regions are linked to the establishment of hearing organs in fish, as these precursor tissues will give rise to the inner ear and the lateral line organ during later stages of development (Haddon and Lewis 1996; Whitfield et al. 2002). In 72 hpf embryos, the expression of *frmpd4* is restricted to cells of the fore- and midbrain, the eyes and the otic vesicle, while expression in the lateral line was no longer detected at this time point. *frmpd4* expression in the otic placode/otic vesicle is restricted to distinct cells clusters and resemble regions of neuronal differentiation (regions 4 and 5 in Figure 2C) (Kantarci et al. 2015). The *frmpd4* expression domain co-localizes with *neurogenin1* (*neurog1*) at these stages, a key factor in otic neurogenesis (Supplementary Figure 5A) (Andermann et al. 2002). Knockdown of *neurog1* resulted in reduction of *frmpd4* expression in the otic vesicle and thereby indicate partial regulation of *frmpd4* by *neurog1* (Supplementary Figure 5B), while Morpholino knockdown of *frmpd4* did not result in prominent changes of *neurog1* or *isl1* expression patterns (Supplementary Figure 5C and 5D).

**>Supplementary Figure 5**

In summary, non-syndromic hearing loss and FRMPD4 expression patterns in hearing organs of vertebrates strongly implied a conserved molecular function during this process. To further investigate this, we conducted a study of hearing capacity in *frmpd4* loss of function models.

### FRMPD4 ortholog Ngh (CG42788) is required for proper hearing in *Drosophila melanogaster*

In *Drosophila melanogaster*, the gene *CG42788* (Supplementary Figure 6), which we renamed to *nicht gut hörend (ngh)*, is the sole ortholog of vertebrate *frmpd4* (28% sequence identity and 44% sequence similarity). RNA levels in KO mutant flies revealed practically no residual expression of *ngh* (Figure 3A). The mutants were viable, and the gross morphology of their antennal ears appeared to be unaffected by the loss of *ngh*. Initial database screening of the *CG42788* (*ngh*) gene imply a rather broad expression pattern during developmental stages and in adult organs, e.g. adipose, reproductive and muscle system. Specific reporter gene analysis, using a hexameric cytoplasmic GFP and a *ngh^1,Gal4>^ promotor trap*, revealed a weak expression in Johnston’s organ neurons (JONs), the chordotonal mechanosensory neurons that mediate hearing in the fly (Albert and Gopfert 2015) (Figure 3B). To assess auditory function, we first measured the mechanical free fluctuations of the antennal sound receiver in the absence of acoustic stimuli. These free fluctuations are actuated by thermal bombardment and motile responses of JONs (Göpfert et al. 2005). Compared to the antennal receivers of controls, those of *ngh* KO flies fluctuated with a lower power (Figure 3C), signaling a reduction of JON motility. The fluctuations were tuned to the same resonant frequency in mutants and controls (242 and 232 Hz, respectively), and we next stimulated the flies with pure tones at that frequency, while varying the sound particle velocity (Göpfert et al. 2006). When we plotted the displacement of the antennal receiver against the particle velocity, we found that the compressive nonlinearity is more pronounced in controls than in *ngh* KO flies. This compressive nonlinearity reports mechanical amplification by JON motility, which nonlinearly boosts the mechanical sensitivity of the antennal receiver when sound is faint (Göpfert et al. 2006). Compared to controls, the gain of this mechanical amplification was reduced in *ngh* KO flies, signaling that loss of *ngh* impairs JON motility and active mechanical amplification in the ear. Along with these mechanical defects, slightly, though non-significantly larger sound particle velocities were required in controls to evoke compound action potentials of JONs in the mutants than in controls, and maximum CAP amplitudes were lightly reduced. Hence, hearing in *Drosophila* is modulated by *ngh*, the *Drosophila* ortholog of *frmpd4*.

**>Supplementary Figure 6**

### *frmpd4* KO in zebrafish causes inner ear and posterior lateral line defects

Expression of *frmpd4* in auditory organs and their precursor structures in zebrafish embryos imply a functional conservation or a comparable role to *FRMPD4* during hearing perception in humans. Sound perception in fish is different to hearing in higher vertebrates as anatomical adaptations to water habitats results in the detection of water vibrations and flow changes rather than sound waves (Nicolson 2005; Whitfield 2002). Specialized sensory cell clusters, e.g. neuromast cells of the lateral line organ, are thought to be orthologue structures to hair cells in the cochlea of higher vertebrates and function as mechanoreceptors of the auditory system (Atkinson et al. 2015; Dambly-Chaudière et al. 2003).

Investigations of *frmpd4*’s role during the development and function of hearing organs in zebrafish were performed by loss of function experiments (different zebrafish genetic tools are summarized in Supplementary Figure 7). We utilized splice site blocking Morpholinos (Supplementary Figure 8), transient F0 CRISPants (Supplementary Figure 9) and splice site deficient *frmpd4* ENU mutants (genomic feature: sa12377; Supplementary Figure 10) to determine the appearance of sensory neuromast clusters marked by the fluorescent vital dye DASPEI (2-(4-(dimethylamino)styryl)-N-ethylpyridinium iodide) (Harris et al. 2003). The *frmpd4*^sa12377^ variant leads to cDNA alterations, stabilized intron 10-11 and results in mis-splicing events (Supplementary Figure 7C), thereby disrupting normal Frmpd4 protein function. In general, reduction of *frmpd4* function resulted in reduction of detectable neuromast clusters in the PLL and in the otic vesicle (OV; Figure 4A). Along with the reduced amount of neuromast clusters in the posterior lateral line (Figure 4B) and in the otic vesicle (Figure 4C), the distance between the single clusters was increased (Figure 4E). Especially homozygous *frmpd4* mutants clearly showed these characteristics, with only an average of 6.64 clusters PLL/larvae (52.7% reduction to control) and an average of 2.00 clusters PLL/larvae (28.2% reduction to control) neuromast clusters in the PLL and the OV, respectively. The changes in the PLL resulted in a decrease of neuromast clusters and an increase of mean distance between the clusters of 2.94% average of relative distance between PLL clusters. Overall embryo and larval morphology was not influenced by *frmpd4* loss of function and neuromast clusters in regions of no *frmpd4* expression, like preoptic and supraorbital, did not show notable reduction in *frmpd4*^sa12377^ mutants (Figure 4D). In support of different genetic models, gain-of-function experiments of full-length FRMPD4 and of patient variants via RNA injection into zebrafish embryos resulted in changed neuromast numbers within the PLL, but did not significantly change other neuromast patterns like neuromast number in OV or neuromast distances (Supplementary Figure 10).

**Figure 4.**
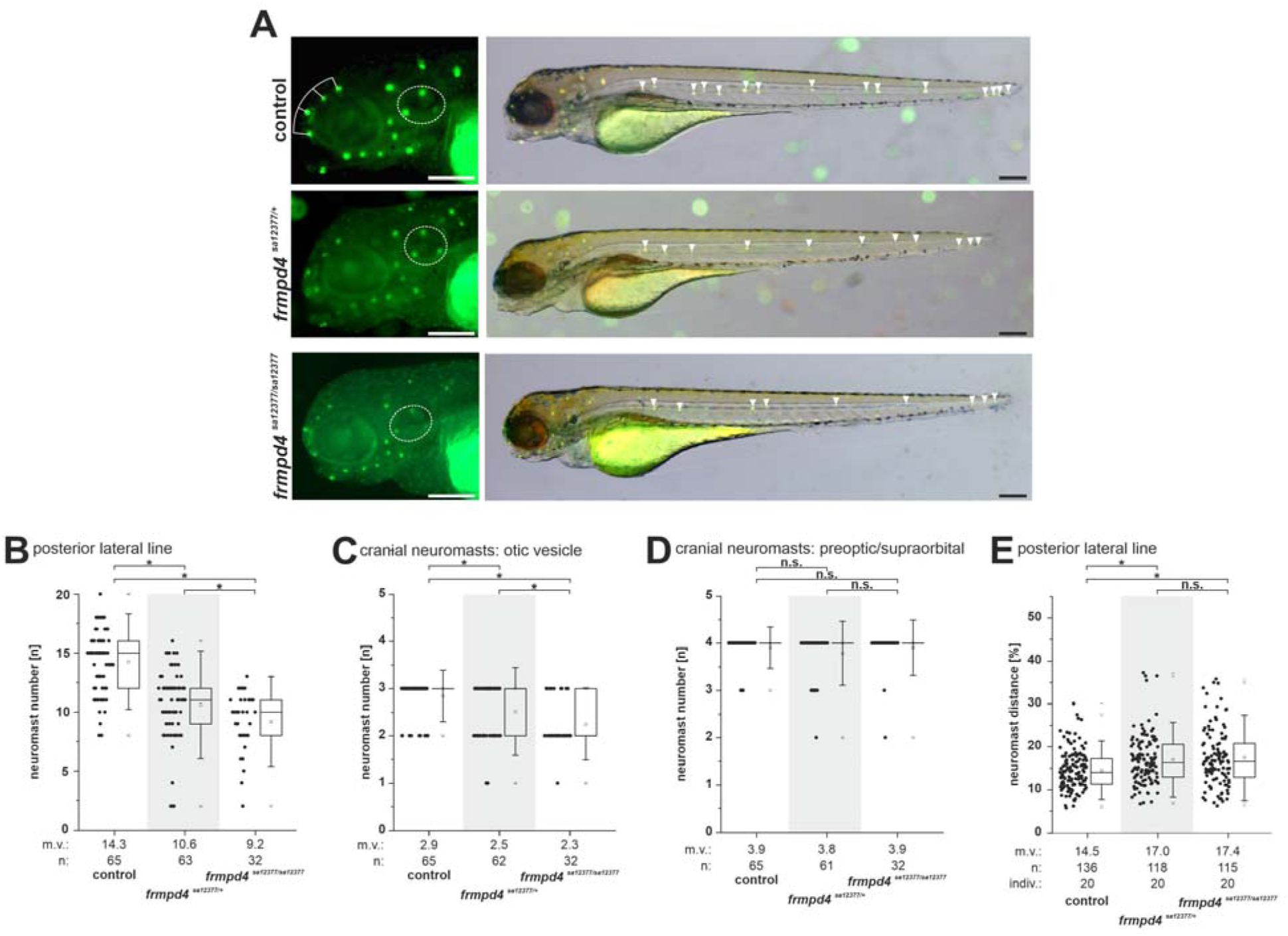
Loss of function of *frmpd4* in zebrafish results in reduced neuromast number and distance changes. **(A)** *frmpd4*^sa12377^ mutants display no general developmental defects, but show loss of DASPEI positive neuromast cells in the posterior otic vesicle (dashed white circle) and in the posterior lateral line (white arrowheads). Quantification of DASPEI positive neuromast cell number in the posterior lateral line **(B)** and the otic vesicle **(C)** showed significantly reduced amounts in heterozygous and even stronger reduction in homozygous *frmpd4* mutants. **(D)** Preoptic and supraorbital neuromast cells are not lost in the *frmpd4*^sa12377^ mutants (the corresponding cell cluster dorsal of the eye marked with white lines in A). **(E)** Measurements of relative distance between neuromasts in the posterior lateral line indicate increased relative distances between clusters in *frmpd4^sa12377^* mutants. m.v.: mean value; n: data point amount; indiv.: number of investigated individuals; statistical significance was calculated by a two-tailed Mann-Whitney U test. An asterisk indicates significant changes between groups, while n.s. marks not significantly different groups. Scale bars in A indicate 100µm.

**Figure 5.**
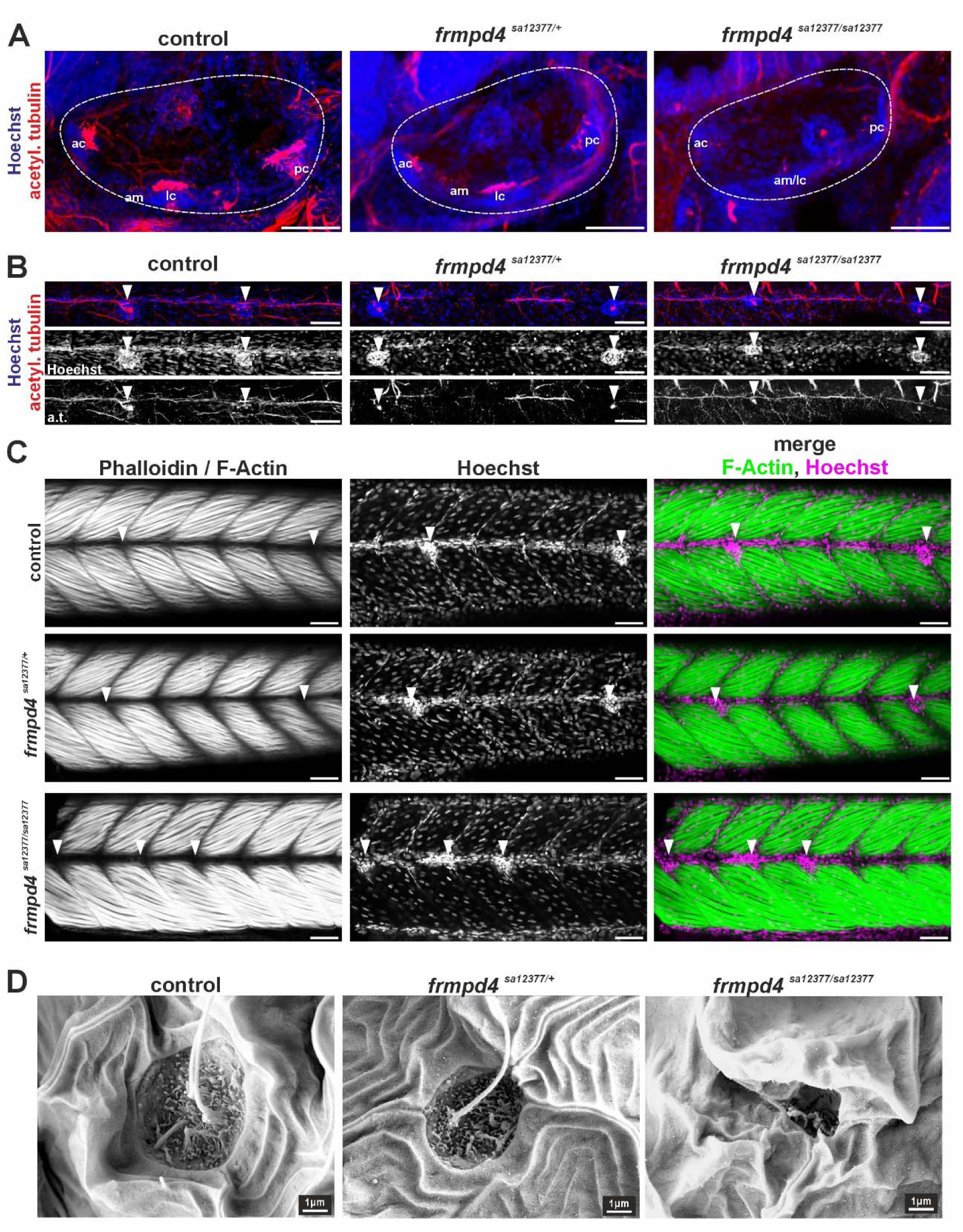
Loss of function of *frmpd4* in zebrafish results in axonal and structural malformations in the otic vesicle and the posterior lateral line. **(A)** Staining for acetylated tubulin in 4 dpf embryos indicated loss of neuronal cell and axonal projection in ventral sensory patches of the otic vesicle in homozygous *frmpd4^sa12377/sa12377^* mutants. **(B)** Posterior lateral line neuromasts and axons (white arrowheads) are affected by *frmpd4* loss by depicting size reduction and morphological changes (analyzed embryos per genotype: 7 wild type controls; 6 heterozygous *frmpd4^sa12377/+^*; 5 homozygous *frmpd4^sa12377/sa12377^*). **(C)** Neuromast cell deposition in the PLL can be disrupted in homozygous *frmpd4^sa12377/sa12377^* mutants, while adjacent somites show normal patterns (n=3 embryos per genotype; control and heterozygous *frmpd4^sa12377/+^* mutants display indistinguishable patterns). **(D)** Scanning electron microscopy further showed disruption of cellular organization in PLL neuromasts and loss of kinocilia in in *frmpd4^sa12377/sa12377^* mutants (n=3 per genotype). m: anterior macula, pm: posterior macula, ac: anterior crista, lc: lateral crista, pc: posterior crista. Scale bars in A to C indicate 50 µm.

**>Supplementary Figures 7-10**

To further investigate the neuronal and structural consequences of *frmpd4* loss on sensory patches in the OV and in the PLL, we performed immunofluorescence staining of acetylated tubulin in axons of 4 days post-fertilization (dpf) old fish larvae (Figure 5A and B; Supplementary Figure 8F and 8G). In the OV, this staining marks sensory patches corresponding to pseudostratified epithelium of the inner ear consisting of sensory hair cells and supporting cells, like the anterior and posterior macula, the anterior, posterior and lateral cristae (Haddon and Lewis 1996). Reduction of Frmpd4 function in heterozygous *frmpd4^sa12377/+^* and most prominently in homozygous *frmpd4^sa12377/sa12377^*mutants results in reduction of neuronal cell and axonal projection in these sensory patches, especially in the posterior cristae (Figure 5A). Neuromasts are deposited along the PLL and can be identified by their typical structure visible by nuclear staining. In accordance with the otic vesicle phenotype and the DASPEI staining (Figure 5B), PLL neuromasts are affected in *frmpd4^sa12377/sa12377^* mutants. They depict reduced axonal outgrowth, reduction of cell nuclei in neuromasts and potentially a reduced lateral line nerve, as indicated by weaker acetylated tubulin signal (Figure 5B). To exclude structural changes caused by defects in surrounding somite muscles tissues, F-actin/Phalloidin staining was performed (Figure 5C). This experiment showed normal muscle development in controls and in *frmpd4^sa12377/sa12377^* mutants, although localization of neuromasts is altered (indicated by white arrowheads). To investigate potential structural or anatomical changes in PLL neuromasts, we performed high magnification scanning electron microscopy (Figure 5D). While control or *frmpd4^sa12377/+^* heterozygous individuals did not show severe morphological disruptions in PLL neuromasts, *frmpd4^sa12377/sa12377^* individuals depict prominent disrupted structures, with reduced kinocilia and collapsed epidermal openings. Taken together, these results suggest a loss of hearing capacity in zebrafish after loss of Frmpd4 function due to either neurological or anatomical changes.

### Loss of Frmpd4 function in zebrafish results in reduced reactions to acoustic stimuli

Our zebrafish investigations indicated neuronal and anatomical alterations in zebrafish larvae and implied influence on sound perception and subsequently on behavior patterns. Normal zebrafish sound perception is fast and can result in an autonomous reflex called the “startle response”. This response represents an instinctive escape behavior to sudden, unexpected threats and stimuli (Colwill and Creton 2011; Zeddies and Fay 2005). We tested adult *frmpd4^sa12377^* mutants in a hearing set up and quantified the appearance of startle responses and the response time after a given sound stimulus (Figure 5A; Supplementary Movie 1 and 2). WT and animals heterozygous for the *frmpd4^sa12377^*allele reacted similarly to a given sound stimulus, while animals homozygous for the *frmpd4^sa12377^*allele reacted at a much lower frequency (Figure 5B). Measurement of the reaction time further indicated that animals of the *frmpd4^sa12377/sa12377^* group also reacted much slower to a given sound stimulus (Figure 5C).

**Figure 6.**
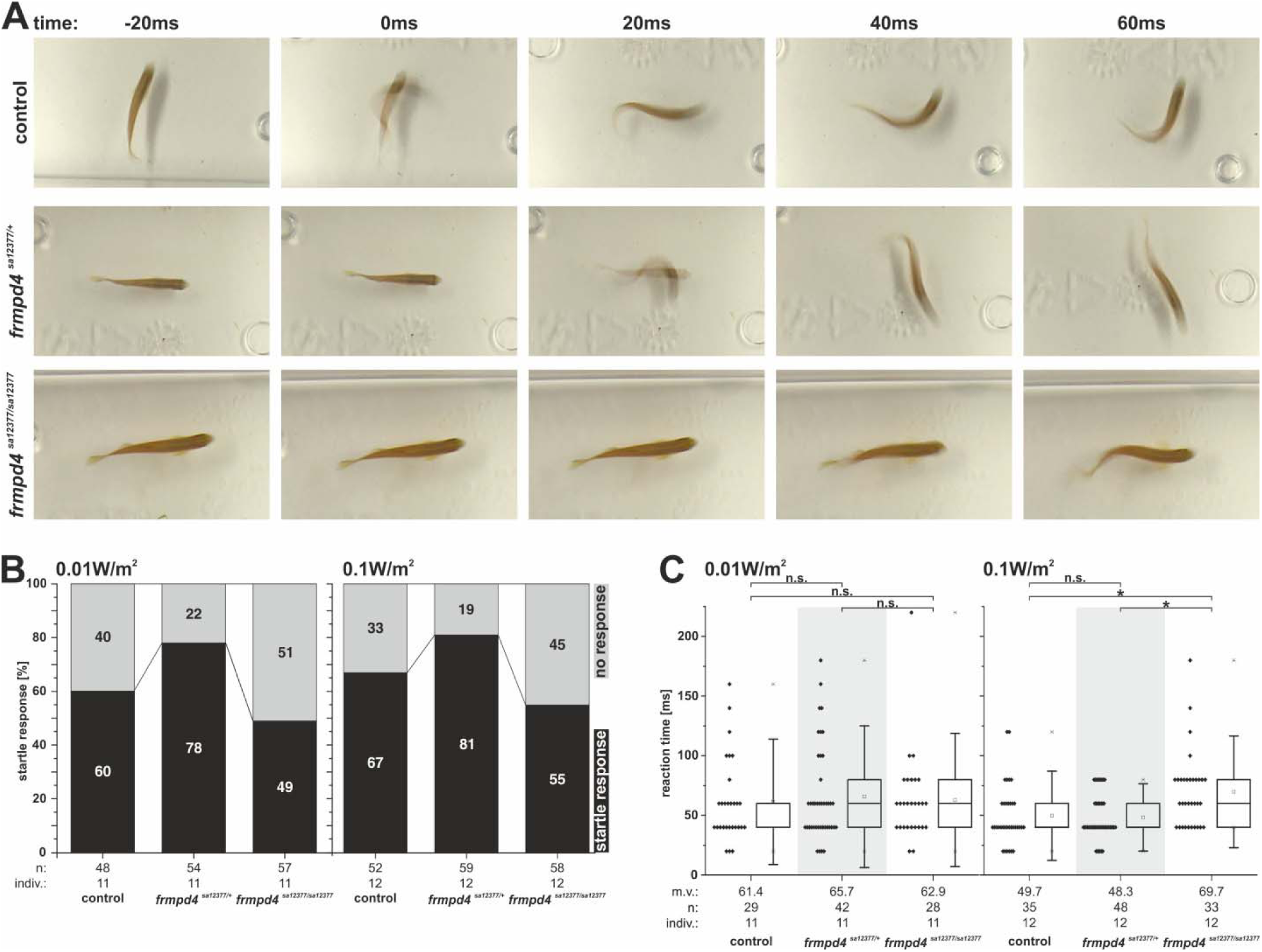
Loss of function of *frmpd4* in zebrafish results in reduced startle response behavior and slower reaction times. Startle response reaction to a given sound stimulus (sound levels in air low dB: 100 dB = 0.01 W/m^2^; high dB: >110 dB = 0.1 W/m^2^; 4400 Herz, 20 ms) is hampered in *frmpd4^sa12377/sa12377^* mutants. **(A)** Single images from high-speed recordings of fishes reacting to a given sound stimulus (0ms). Quantification of startle responses **(B)** and reaction times **(C)** indicate reduced acoustic perception and slower response in *frmpd4^sa12377/sa12377^* mutants. m.v.: mean value; n: data point amount; indiv.: number of investigated individuals. Statistical significance as calculated by a two-tailed Mann-Whitney U test. Asterisks indicate significant changes between groups, while n.s. marks not significantly different groups. Whiskers indicate standard deviations (coefficient: 1.5).

### *Frmpd4^-/-^* mice have reduced ABR amplitudes and high-frequency hearing loss

Cochlear morphology was investigated in conditional KO mice, originally called *Preso1^-/-^*in initial studies, hereafter referred to as *Frmpd4^-/-^* mice, where they exhibited sustained, metabotropic glutamate receptor (mGluR) 5-dependent inflammatory pain linked to enhanced mGluR signaling (Hu et al. 2012). Although these mice were viable, fertile, and showed similar development and breeding behaviors as WT littermates, the cochlear epithelium of *Frmpd4^-/-^* KO animals was morphologically altered in histological sections at level of the outer and inner hair cells visualized by F-Actin/Phalloidin localisation (Figure 6A). Simultaneously, unchanged TUBB3 expression was detectable in WT and KO animals and implies normal neuronal development in the organ of Corti. Subsequent functional measurements of hearing capacity imply that waveforms of *Frmpd4^-/-^* are smaller with shifts in latency (Figure 6B). Individual (Figure 6C) and mean (Figure 6D) ABR thresholds with clicks and tones indicated high frequency hearing loss of *Frmpd4^-/-^*mice, with significant differences seen at 24 (t(49)=11.6, p <0.0001) and 32 kHz (t(49)=3.2, p= 0.0134), but not other frequencies. In the mixed model, the interaction between genotype and frequency was significant (F(6,48)=17.2, p<0.0001), reflecting the observation that hearing loss in *Frmpd4^-/-^* mice was limited to high frequencies.

**Figure 7.**
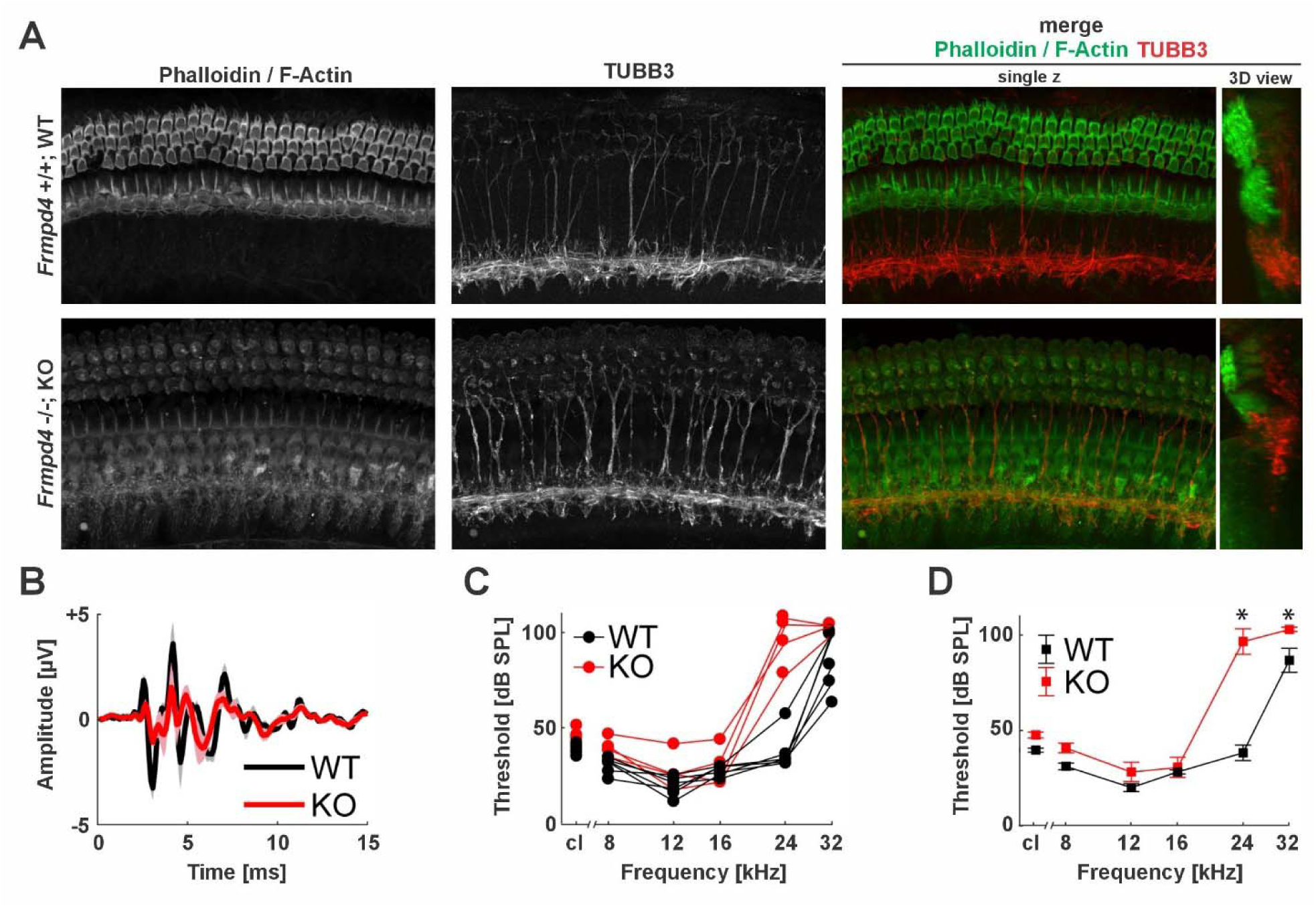
Knockout of *Frmpd4* displays morphological changes in the cochlea and elevates high-frequency ABR thresholds. **(A)** Confocal laser scanning images of mouse cochlear display altered epithelial structures in *Frmpd4* KO, visualized by top-down views and simultaneous Phalloidin (F-Actin) and TubB3 staining. **(B)** Grand average ABR waveforms in response to a click presented at 90 dB peSPL. Shaded regions indicate standard deviation of the mean. **(C)** Individual ABR thresholds in response to clicks (cl) and tones. **(D)** Mean ABR thresholds in response to clicks and tones. Error bars represent standard error of the mean. Asterisks indicate significant differences between genotypes.

**Figure 8.**
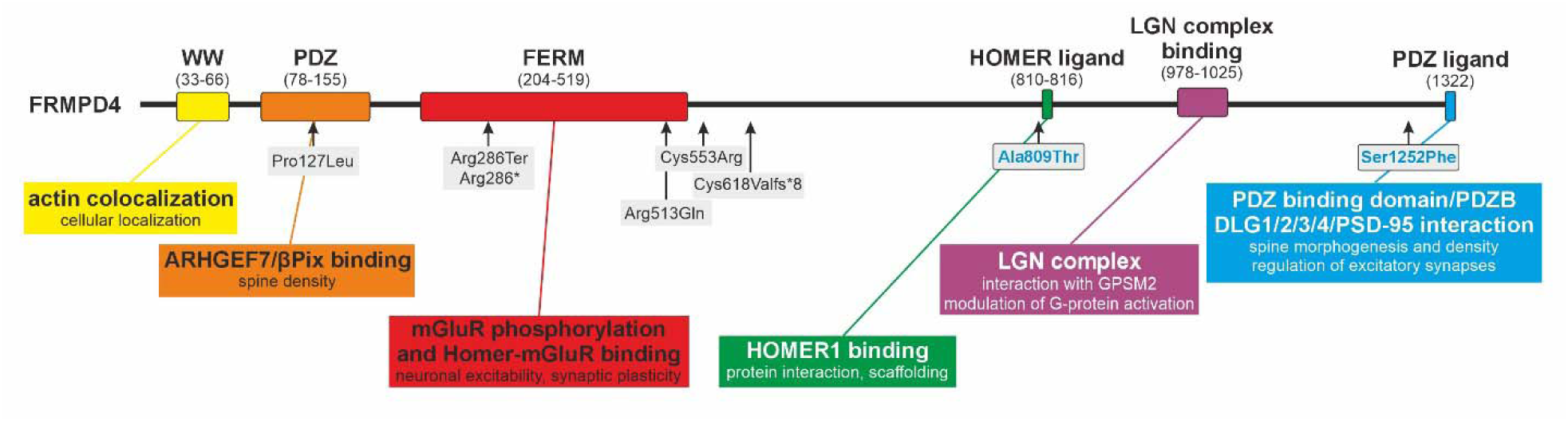
Overview of described FRMPD4 molecular functions linked to previously described variants and known protein domains. Newly reported genetic variants described in this study are located at the C-terminus and are highlighted in boxes with blue writing.

## Discussion

The identification of genetic causes of non-syndromic hearing loss is important for accurate diagnosis, prognosis, and clinical management. We describe two families with maternally inherited *FRMPD4* missense variants with bilateral sensorineural non-syndromic hearing loss. Intellectual disability and other phenotypes such as epilepsy that have been attributed to *FRMPD4* variants were not observed in these individuals. Extensive annual or semi-annual medical follow-up of Family 1 for more than 20 years suggests that a developmental phenotype is unlikely. This work expands the phenotypic spectrum associated with disease-associated variants in *FRMPD4* and has implications for clinical management, molecular genetic diagnostic testing, genetic counseling, therapeutic decision-making, while also opening new avenues for studying mutational mechanisms and underlying pathophysiological processes.

To investigate molecular mechanisms linking *FRMPD4* to hearing loss, we analyzed its expression in auditory organs across several vertebrate species. Investigation of *Frmpd4* expression and protein localization in mouse and rat cochleae indicated general expression in neuronal tissues, with comparatively low expression levels in the cochlea. Further analysis of RNA sequencing data from the developing mouse cochlea showed evidence of increasing expression in the spiral ganglion neurons that is sustained into adulthood.

Subsequent protein localization further clarified specific FRMPD4 localization in organ of Corti and spiral ganglion during development (stage P1–P28), partly resembling the general spiral ganglion neuron marker TUBB3. Investigation of *frmpd4* expression in zebrafish embryos showed transcripts in early progenitor cells of corresponding hearing organs (OV and PLL).

Although expression in zebrafish embryos is highly dynamic, sustained expression was identified in the OV in a pattern partly localized with *neurog1*, a marker for early neural differentiation (Korzh et al. 1998) and cranial sensory ganglia (Andermann et al. 2002; Yu et al. 2020).

The association between FRMPD4 and hearing appears to be evolutionarily conserved in *Drosophila*, where the FRMPD4 ortholog *ngh* is expressed in JONs, the auditory sensory cells of the fly. Loss of the *ngh* function impairs JON motility, resulting in reduced mechanical amplification during hearing, and decreased maximum amplitudes of sound-evoked CAPs of the JONs. The residual mechanical amplification and CAPs that persist in *ngh* KO flies imply that only a subset of the approximately 500 JONs in the *Drosophila* ear are functionally compromised in the KO flies, despite apparent *ngh* expression across all JONs. Follow-up studies will be required to clarify how *ngh* dosage contributes to JON function and to further assess conservation of auditory mechanisms across species.

Loss of function experiments in zebrafish, including splice site deficient ENU mutants, splice site blocking Morphants, and transient CRISPR/Cas9 genetic approaches, indicate a role for *frmpd4* in the development of auditory organs during embryonic and early larval development, as shown by reduced neuronal cell populations in both the OV and PLL. Additionally, morphological alterations in PLL neuromasts further support a structural role for Frmpd4 and suggest an influence on neuronal cell number. Migration of PLL primordium cells is not abolished following Frmpd4 reduction, as neuromasts are still detected along the entire body axis of affected embryos. Moreover, preliminary experiments indicate that expression of specific PLL markers *tfap2* and *cldnb* is preserved in *frmpd4* morphants (data not shown). Nevertheless, the reduced number of neuromast clusters, combined with fewer neuronal cells per neuromast is likely to impair function. Given that *frmpd4* expression was detected in the PLL primordium but not in neuromast cells, an early developmental effect of Frmpd4 on the PLL is plausible.

Consistent with these findings, hearing perception in *frmpd4* adult mutants is altered, as the startle response to sound stimulus is reduced and slower. This behavioral observation might be linked to additional *frmpd4* functions during neuronal development and cannot be separated from complex, higher brain function of adult individuals, but this phenotype mirrors a hearing-impaired situation similar to humans. Future studies will be required to determine whether these effects arise due to loss of PLL primordium cells, neuronal malformations, or result from morphological alterations of sensory organs. Recently, direct protein interaction between FRMPD4 and Whirlin-a (a protein associated with Usher syndrome type 2) was reported in the zebrafish retina, providing the first evidence for a potential postsynaptic protein complex involving FRMPD4 (Schellens et al. 2022). These findings hint to a neurological mode of Frmpd4 function mediated by interaction with Whirlin through C-terminal interaction via PDZ-binding motifs) and subsequent modulation of signaling via the GPSM2/LGN complex (Mauriac et al. 2017; Takayanagi et al. 2015; Yuzawa et al. 2011).

Remarkably, the mouse model presented here recapitulates the auditory phenotype observed in humans and argues against the presence of severe neurodevelopmental phenotypes previously associated with *FRMPD4*. Although a *Frmpd4^tm1dIcs^*mouse line was previously characterized by the International Mouse Phenotyping Consortium (IMPC) and reported to have a non-significant ABR, acoustic startle, and pre-pulse inhibition results, these findings suggest that large-scale phenotyping pipelines focusing on young adult animals may lack sensitivity for detecting hearing impairment in older mice, particularly when higher frequencies are severely affected. Secondly, this mouse model was originally studied in the context of inflammatory pain without suspicion of an auditory phenotype (Hu et al. 2012), underscoring the importance of detailed and highly sensitive ABR testing. Behavioral audiogram thresholds may not be good predictors of ABR thresholds as the central auditory pathway can compensate for some peripheral dysfunction (e.g. central gain compensation).

The high-frequency hearing loss observed in *Frmpd4^-/-^*mice is most similar to the phenotype observed in Family 1, where substantial residual hearing is present despite pan-frequency involvement. The comparatively more severe hearing loss in the proband of Family 2 remains unexplained, and we cannot exclude the contribution of genetic modifiers or additional undetected genetic variants contributing to these differences. Variable expressivity is common in hereditary hearing loss, including intrafamilial variability as observed in Family 1, where individual III:3 presented with moderate hearing loss and III:2 with mild hearing loss. Such variability may also have contributed to delayed recognition of hearing loss in the older sibling (III:2), who was diagnosed only retrospectively following newborn hearing screening and clinical diagnosis of his younger sibling.

The patient variants described in this study are located at the C-terminal region of FRMPD4 and do not affect well-characterized functional domains at the N-terminus (see schematic overview in Figure 7). Protein structure prediction tools, such as by AlphaFold (Jumper et al. 2021; Varadi et al. 2022), perform poorly in these evolutionarily less conserved, intrinsically disordered regions and therefore do not provide reliable structural insight (UniProt: Q14CM0). Notably, the individuals reported here do not exhibit intellectual disability or epilepsy, in contrast to previously described pathogenic *FRMPD4* variants, including deletions or and truncating variants (listed in Supplementary Table 7). A previously reported patient with complete loss of the FRMPD4 N-terminus presented with X-linked intellectual disability and a dendritic spine density phenotype was functionally quantified (Piard et al. 2018), indicating a more severe neurological impact. This contrast suggests that the missense variants identified here do not result in complete loss of function, or that different FRMPD4 protein domains contribute to distinct phenotypes.

One hypothesis to be explored in future studies is that these newly associated *FRMPD4* missense variants with hearing loss interfere with neighboring protein domain interactions, leading to hearing loss without broader neurodevelopmental consequences. Candidate interactors include HOMER proteins, which share related functional features and exist in multiple transcript classes (Soloviev et al. 2000). *HOMER1* combines various molecular functions as it acts, on the one hand, as a scaffolding/multimodal adaptor protein (reviewed in (de Bartolomeis et al. 2022)). It further displays protein multimerization with CDC42 (Shiraishi-Yamaguchi et al. 2009), cytoskeletal organization during postsynaptic density to regulate homeostatic synaptic plasticity (Heavner et al. 2021). HOMER2 is associated with autosomal dominant non-syndromic hearing loss (DFNA68) and functional modeling in mice induces stereocilia abnormalities after over-expression and early-onset hearing loss after KO, respectively (Azaiez et al. 2015). We speculate that spatial-temporal interactions of FRMPD4 and HOMER proteins contribute as a key driver of diverse physiological outcomes. An alternative mechanism may involve FRMPD4 interaction with the LGN/GPSM2 adaptor protein complex via TPR-domain-mediated binding, as previously experimentally resolved for FRMPD1 (Takayanagi et al. 2015; Yuzawa et al. 2011). The LGN/GPSM2 complex is implicated in diverse cellular processes, including symmetric cell division, spindle orientation/localization, and neural stem cell division in the neuroepithelium (Bhonker et al. 2016; Konno et al. 2008; Mauriac et al. 2017). Disruptions of these pathways have been linked to hearing impairment through effects on actin-rich stereocilia elongation in auditory and vestibular hair cells, stereocilia row assembly, and regulation of actin dynamics in epithelial and neuronal tissues (Mauriac et al. 2017; Shi et al. 2022; Tadenev et al. 2019). Finally, the N-terminal PDZ domain of FRMPD4 has been shown to regulate dendritic spine morphology via interaction with PSD-95 (Lee et al. 2008) but has not yet been linked to hearing impairment. Nonetheless, PDZ domain-containing proteins are repeatedly implicated in auditory disorders, including Usher syndrome, where PDZ domain-containing scaffolding proteins such as WHRN (USH2D or DFNB31) orchestrate assembly of the USH2 complex (Stemerdink et al. 2022). Taken together, we speculate that the newly identified *FRMPD4* variants result in slight modulation of protein-protein interactions involving HOMER proteins, the LGN/GPSM2 complex, or PDZ-domain interactors, leading specifically to non-syndromic sensorineural hearing loss without broader neurodevelopmental manifestations.

## Conclusions

In summary, we identify *FRMPD4* as a novel X-linked gene for non-syndromic sensorineural hearing loss, expanding its previously described phenotypic spectrum beyond intellectual disability and epilepsy. Cross-species functional analyses in *Drosophila*, zebrafish, and mouse collectively establish an evolutionarily conserved role for FRMPD4 in auditory function, while the absence of neurodevelopmental features in affected individuals suggests that distinct protein domains and interaction networks underlie its diverse phenotypic manifestations. These findings have direct implications for molecular genetic diagnostics and variant interpretation in patients with hearing loss, and implicate *FRMPD4*-mediated interactions with HOMER proteins, the LGN/GPSM2 complex, and PDZ-domain partners as candidate mechanisms warranting investigation in future studies.

## List of abbreviations

ABR: auditory brainstem response
ax: axon
cd: ciliary dilation
ci: cilium
cl: clicks
DASPEI: 2-(4-(dimethylamino)styryl)-N-ethylpyridinium iodide
de: dendrite
dpf: days post-fertilization
*FRMPD4*: FERM and PDZ Domain Containing 4
gDNA: genomic DNA
HL: hearing level
hpf: hours post-fertilization
IMPC: International Mouse Phenotyping Consortium
indiv: number of investigated individuals
JO: Johnston’s organ
JONs: Johnston’s organ neurons
KO: knockout
m.v.: mean value
mGluR: metabotropic glutamate receptor
n: data point amount
ngh: nicht gut hörend
n.s.: not significant
OV: otic vesicle
PLL: posterior lateral line
PTA: pure-tone average
so: soma
WT: wild type

## Declarations

### Ethics approval and consent to participate

The ethics commission of the Medical Faculty of the University of Würzburg, Germany, gave ethical approval for this work (approval number 46/15).

All procedures involving experimental animals were performed in compliance with local animal welfare laws (Tierschutzgesetz §11, Abs. 1, Nr. 1), European Union animal welfare guidelines (EU directive 2010/63/EU), Johns Hopkins School of Medicine Institutional Animal Care and Use Committee (IACUC), and best-practice scientific policies (ARRIVE guidelines, the *Guide for Care and Use of Laboratory Animals*). Presented zebrafish embryo and larvae experiments have been performed at stages younger than 5 dpf and have been terminated timely before free-swimming and independent feeding larval stages. Experiments including fin clip genotyping and adult startle response imaging were officially permitted by the local authorities (permit number: DMS-2532-2-9 and DMS-2532-2-428). Procedures involving transgenic animals were in addition performed according to local regulations on this topic (Gentechnikrecht/Gentechnik-Sicherheitsverordnung).

## Consent for publication

Consent has been obtained for publication.

## Availability of data and materials

In addition to the supplementary files, essential raw data files linked to this paper have been deposited at the Zenodo digital repository and are freely available for download here: **10.5281/zenodo.18507823**

The repository data includes quantification tables for RT-qPCR experiments, quantification tables for zebrafish DASPEI experiments, mouse and *Drosophila* hearing measurements, additional images not used for figures and overview figures, specification of zebrafish hearing set-up and high-speed movies used for startle response quantification.

Additional data supporting this study’s findings are available from the corresponding author upon reasonable request.

## Authors’ contributions

Conceptualization: D.L., T.H., B.V.; Data curation: D.L., K.R., K.M.S., R.K., N.C., D.B., S.H., J.D., P.R.S., L.B., J.V. C.N., M.A.H.H., J.S., W.S.D., M.C.G., A.M.L., B.V.; Formal Analysis: D.L., K.R., K.M.S., R.K., N.C., D.B., S.H., J.D., I.N., J.D., P.R.S., L.B., C.N., M.A.H.H., J.S., W.S.D., M.C.G., A.M.L., B.V.; Funding acquisition: D.L., T.H., B.V.; Investigation and validation: D.L., K.R., K.M.S., R.K., N.C., D.B., S.H., J.D., I.N., J.D., P.R.S., L.B., C.N., M.A.H.H., J.S., R.T.W.S, E.d.V, E.v.W, W.S.D., M.C.G., A.M.L., B.V.; Methodology: D.L., K.R., K.M.S., R.K., N.C., D.B., S.H., J.D., I.N., M.C.G., A.M.L., L.B., M.C.G., H.G., A.M.L., T.H., B.V.; Project administration: D.L., K.R., K.M.S., M.C.G., A.M.L., B.V.; Resources: U.Z., O.B., E.K., W.S.D., R.M., T.W., P.F.W., M.C.G., H.G.; Software: S.H., P.H., T.M., M.D., P.M.K., R.M.; Supervision: I.N., E.K., M.C.G., H.G., T.H.; Writing – original draft: D.L., B.V.; Writing – review and editing: all authors have read and approved of this work.

## Data Availability

In addition to the supplementary files, essential raw data files linked to this paper have been deposited at the Zenodo digital repository and are freely available for download here: 10.5281/zenodo.18507823

## Acknowledgements

The authors are grateful to the families for their participation. We thank Helmut Korder, Laura Halbhuber, and Johannes Volker for technical support. We are grateful to Kristina Luermann and Marcus Müller for productive discussions with the *Frmpd4* measurements and to Dr. Claudia Davenport for critical reading. Zebrafish ENU mutant lines were kindly provided by the European Zebrafish Resource Center (EZRC, KIT Karlsruhe, Germany). We thank Christian Stigloher, Claudia Gehrig and Daniela Bunsen from the Electron Microscopy Core Unit of the Biocenter at the University of Würzburg, for support with the electron microscopy analysis. This work was done with the support of the Center for Rare Hearing Disorders at the Center of Rare Diseases Göttingen (ZSEG). We are indebted to all our colleagues for fruitful scientific discussions.

## Supplemental Materials and Methods

### Clinical evaluation and human genetics methods

#### Clinical assessment

Diagnosis of sensorineural hearing loss was achieved according to current clinical standards and applied age-appropriate methods to determine hearing thresholds at routinely measured frequencies. Hearing loss threshold descriptions followed GENDEAF standards (Mazzoli et al. 2003). Routine pure-tone audiometry was performed on individuals III:2 and III:3 in Family 1 and III:2, and IV:1 of Family 2 according to current standards and measured hearing thresholds at 0.25, 0.5, 1, 2, 4, and 8 kHz. Air- and bone-conduction thresholds were measured and severity of hearing loss was determined by averaging pure-tone thresholds over 0.5, 1, 2 and 4 kHz (pure-tone average, PTA_0.5-4K_). Otoacoustic emissions neonatal hearing screening and auditory brainstem response (ABR) testing was performed in individual III:3 of Family 1. Transient evoked otoacoustic emissions and distortion products were further tested in III:2, III:3 (Family 1) and IV:1 (Family 2). Auditory steady-state response was performed in IV:1 (Family 2). Tympanometry was performed in the affected individuals in Family 2. The severity of hearing loss in the better ear was defined as mild for thresholds averaging 20-40 dB hearing level, moderate for 41-70 dB hearing level, severe for 71-95 dB hearing level, and profound in excess of 95 dB hearing level. Progressive hearing loss was defined as a deterioration of >15 dB hearing level in the average over the frequencies of 0.5, 1, and 2 kHz within a 10-year period.

#### Exome data filtering, analysis, and variant prioritization

Exome data were filtered through pedigree-based strategies to evaluate autosomal recessive, autosomal dominant, and X-linked modes of inheritance. The bioinformatics filtering strategy focused on exonic and donor/acceptor splicing variants as previously described (Vona et al. 2021). Alternative alleles present at >20% and a minor allele frequency <0.01 were assessed using gnomAD v4.1.0 (Chen et al. 2024), TopMed freeze.8 (Taliun et al. 2021), and data from the All of Us research program (The All of Us Research Program Genomics Investigators et al. 2024). MAF thresholds were later adjusted according to ACMG/AMP guidelines for hearing loss (Oza et al. 2018) and an in-house epilepsy cohort (n=511) with a threshold of ≤1%. Artifact-prone gene families (*HLA*s, *MAGE*s, *MUC*s, *NBPF*s, *OR*s, *PRAME*s) were excluded. Variant types that were analyzed included SNVs, indels, and splicing variants with potential structural or cryptic splice site effects. Variants in 5’ and 3’ UTRs, deep intronic, and intergenic regions were excluded. Allele read frequencies between 25-75% were classified as heterozygous; those >75% as homozygous. Clinical databases ClinVar (Landrum et al. 2025) and the Deafness Variation Database (DVD) (Azaiez et al. 2018) were referenced to assess already reported interpretations.

Pathogenicity of variants was assessed using multiple *in silico* tools: SIFT (Schwarz et al. 2014), PolyPhen-2 (Adzhubei et al. 2010), FATHMM (Shihab et al. 2013), MutationTaster (Steinhaus et al. 2021), REVEL (Ioannidis et al. 2016), ClinPred (Alirezaie et al. 2018), and CADD (Schubach et al. 2024). Computational assessment of splicing effects used SpliceSiteFinder-like, MaxEntScan, NNSplice, and GeneSplicer embedded in Alamut Visual Plus v1.12 (Sophia Genetics, Bidart, France) as well as SpliceAI Visual and AbSplice embedded in SpliceAI Visual (De Sainte Agathe et al. 2023). Residual Variation Intolerance Score (RVIS) was used to assess gene intolerance to functional variants (Petrovski et al. 2013).

### Drosophila methods

#### Animal models

Fly (*Drosophila melanogaster*) stocks used in this study and their respective origin were *ngh^MI02203^* (BDSC 60756) mutants and *w^1118^*(BDSC 3605) controls. For expression analysis, the Gal4/UAS-system (Brand and Perrimon 1993) was utilized, with *dnai2-Gal4* (Karak et al. 2015) and hexameric GFP (BDSC 52261).

Targeted deletion and Gal4 knock-in at the *CG42788* (*ngh*) locus were achieved using CRISPR/Cas9-mediated genome editing. Guide RNAs were designed with the Target Finder tool (https://flycrispr.org/) and cloned into the vectors pBFv-U6.2 (Addgene #138400) and pBFv-U6.2B (Addgene #138401). Both gRNA cassettes were subsequently combined into a single construct via *NotI* and *XhoI* restriction sites. For the donor construct, a 1032 bp fragment upstream of the *ngh* transcriptional start site was cloned into the phD-DsRed vector (Addgene #51434), followed by insertion of the *Gal4* coding sequence derived from the pt-Gal4 plasmid (Sharma et al. 2002). Downstream of the 3×P3-DsRed selection marker, an 843 bp 3′ homology arm corresponding to the *ngh* locus was inserted, yielding the final plasmid phD-DsRed-*ngh*^upstream^-Gal4-3×P3>DsRed-*ngh*^Downstream^. The donor and gRNA plasmids were co-injected into embryos of *vas-Cas9*(II) flies (BDSC #56552). Correct deletion and targeted integration of *Gal4* at the *ngh* transcription start site were verified by DsRed reporter fluorescence in the eyes and confirmed by sequencing of the edited genomic locus.

Flies were kept at 25°C, 60% humidity on standard cornmeal-yeast medium in a 12-h/12-h light/dark cycle. Experiments were performed in accordance with German Federal regulations (license Gen.Az 501.40611/0166/501).

#### RT-PCR

Three independent cohorts, each consisting of 20 fly heads per genotype, were snap-frozen in liquid nitrogen for RNA extraction. Total RNA was isolated using the ZR Tissue & Insect RNA MicroPrep Kit (Zymo Research Europe GmbH, Freiburg, Germany; #R2030). 1 µg of RNA per sample was reverse-transcribed into cDNA using the QuantiTect Reverse Transcription Kit (Qiagen, Valencia, CA, USA; Cat. No. 205311). For amplification, 10 ng of cDNA was used per reaction (technical triplicates) with the Phire Tissue Direct PCR Master Mix (Thermo Fisher Scientific; F170S) and *ngh*-specific primers, using *Act5C* as the reference gene. Cycling conditions were: 95°C for 5 min, followed by 40 cycles of 95°C for 15 s, 54°C for 30 s, and 72°C for 30 s, with a final extension at 72°C for 10 min. PCR products were separated by agarose gel electrophoresis, stained with ROTI®GelStain (Carl Roth), and visualized using an iBright gel documentation system (Thermo Fisher Scientific). Band intensities were quantified using the iBright analysis software.

#### Immunoflourescence

GFP expression was visualized in fixed adult Johnston’s organ (JO) tissue sections. Staining was performed as described (Hehlert et al. 2025). In brief, fly 2^nd^ antennal segment sections were prepared from five-day adult fly heads. Heads of adult flies, five days post-eclosure, were isolated and fixed in 4% PFA (Merck) for 1 h at room temperature. After a brief wash with 1x PBS (Merck, P4417) heads were embedded in albumiSn gelatine and fixed with 6% PFA in 0.3% PBS-T (Triton X100, Sigma-Aldrich), pH 7.4, at 4°C overnight. The embedded samples were then fixed in fresh 100% methanol (Merck) for 10 min at 4°C and quickly transferred to 1x PBS for re-hydration. 40 µm tissue sections were generated with a microtome, and tissue slices were stored in 1x PBS-T. Blocking was done in blocking buffer (1x PBS-T, 5% normal goat serum, 2% BSA) for 1h at room temperature. Tissues were then incubated with FluoTag®-X4 Atto 488nm anti-GFP (1:1000, NanoTag Biotechnologies, N0304) and Cy3-conjugated goat anti-HRP (1:300, Jackson ImmunoResearch, 123-165-021) in blocking buffer for 2 h at room temperature. Afterwards samples were washed three times in 1x PBS-T (10min, room temperature). Tissue samples were then mounted on microscope slides in DABCO (Carl Roth, 0718.1). Stainings were analyzed with a Leica SP8 microscope (at 20°C) in 8-bit mode using a C-Apochromat 63x/1.40 W Korr FCS M27 objective and the Leica X software. For fluorescence detection, the following settings were used: Atto488 / Alexa-488 (Ex: 488nm; Em: 490–540 nm); Cy3 (Ex: 561nm; Em: 566–600 nm). Images were subsequently processed in Image J v1.49m (NIH) and arranged in Adobe Illustrator CC.

#### Electrophysiological and mechanical recordings of the *Drosophila* ear

Recordings of antennal mechanics and Johnston’s organ neurons (JON) compound action potentials (CAPs) were performed as described (Senthilan et al. 2012). Single flies were completely immobilized ventral side down in paraffin:beeswax (50:50), leaving only the recorded antenna and arista unobstructed. Antennal vibrations were measured at the arista tip using a laser Doppler vibrometer (PSV-400, Polytec, Waldbronn). Acoustic pure tone stimuli were broadcast with a loudspeaker (Visaton W130S) positioned 10 cm behind the animal. Sound particle velocity at the position of the fly was monitored with a pressure-gradient microphone (Emkay NR3158; Knowles Electronics).

Microphone, laser, and electrode signals were simultaneously digitized at 8.2 kHz. Recordings were divided into 1 s segments (rectangular window function) and subjected to discrete Fast Fourier transformation (FFT). 40-70 segments were averaged for determining power spectra of the mechanical fluctuations of the antennal sound receiver in the absence of acoustic stimulation, and ten segments were averaged to assess pure tone-induced responses of the antenna and the JONs.

Power spectra of mechanical free fluctuations were fitted with a Harmonic oscillator function to determine the resonance frequency (best frequency) of the antennal sound receiver (Göpfert et al. 2005). The respective fluctuation power was assessed by integrating the power spectrum for frequencies between 150 and 1,500 Hz. Mechanical sensitivity of the sound receiver was determined from its displacement response to pure tone stimulation at its resonance frequency, by dividing its displacement by the corresponding sound particle velocity. The mechanical amplification gain provided by JON motility was quantified by dividing the mean mechanical sensitivity of the receiver in the low-intensity linear regime by that in the high-intensity linear regime.

During tonal stimulation, Fourier amplitudes of the microphone and laser signals were extracted at the stimulus frequency, whereas CAP amplitudes were quantified at twice that frequency to account for the characteristic frequency doubling of the JO response. Particle velocity (pv) thresholds of the CAPS were determined as the minimum particle velocity required to elicit ≥10% of the normalized CAP response amplitude (Senthilan et al. 2012).

### Zebrafish methods

#### Zebrafish whole-mount *in situ* hybridization

RNA *in situ* hybridization was performed according to standard protocols (Thisse and Thisse 2008). RNA probes were synthesized from cloned partial mRNA sequences (newly cloned *frmpd4* probe included exon 8 to 12 to target FERM domain sequences; *neurog1* (Blader et al. 1997) and *isl1a* (Appel et al. 1995) probes were previously published) of target genes using the DIG or FLU RNA Labeling Kit (Roche). Sense probes were synthesized as a negative control for each anti-sense probe and used under the same reaction conditions. Specific staining was additionally compared to expression patterns deposited in zfin.org. For signal improvement, embryos were incubated after staining in 100% methanol over night at - 20°C prior to rehydration in PBST, mounting and imaging. Embryos were glycerol mounted, viewed either with a Leica S8 APO Stereomicroscope and were photographed using a Leica MC170HD digital camera or a Zeiss Imager A1 and were photographed using a Zeiss MRc5 digital camera. Digital pictures were acquired via the Leica LAS or the Zeiss Axiovision software and arranged using Corel Draw X6 graphics suite.

#### Zebrafish injections, Morpholino knockdown and CRISPR/Cas9 gene editing

One-cell to maximum four-cell stage zebrafish embryos were injected with solutions comprising of an active reagent (e.g. RNA, DNA or Morpholino), Phenol red (pH 7.0; 0.05% final concentration; for visualization of injection solution) and Fluorescein-Isothiocyanate-Dextran (FITC; Sigma-Aldrich; 1 mg/μl; for visualization of successful and uniform injection in 24 hpf embryos). Positively injected embryos were identified 24 hpf by transient green FITC fluorescence and were further analyzed. In general, injection volumes are adjusted to 1/10 volume of the first cell and were controlled by measurements of droplet size in mineral oil on a micrometer scale slide followed by volume calculation (V = 1/6πd3; mean droplet volume: ∼8 nl).

Antisense Morpholino oligonucleotides were synthesized by GeneTools (Philomath, OR, USA). The *frmpd4* splice site Morpholino 5’-TACACCTGTGTGCCACAAAGAGACA-3’ targeted the exon-intron boundary of zebrafish *frmpd4* intron 14-15 and exon 15 (ENSDARE00000872050). The Morpholino is stabilizing an unspliced product with an addition of 330 bp, which is leading to a premature stop codon. The neurog1 Morpholino 5’-ACGATCTCCATTGTTGATAACCTGG-3’ targeted the start codon of *neurog1* and has been previously published (Andermann et al. 2002). A standard control oligo Morpholino 5’-CCTCTTACCTCAGTTACAATTTATA-3’ was injected in equal amounts as a negative control to exclude unspecific effects. Unless otherwise noted, a 0.25 mM Morpholino solution was injected into each embryo.

Injection of two sgRNAs targeting two positions in the zebrafish *frmpd4* gene locus (sgRNA r2 and sgRNA f3; final RNA concentration each 25ng/µl), along with nCas9n RNA (final RNA concentration 150ng/µl) (Jao et al. 2013) results in transient F_0_ CRISPants. Application of this method results in mosaic embryos possessing deletion of a 199 bp gDNA fragment, flanking the conserved Serine position associated with hearing loss in Family 1. Functionality of used sgRNAs and Cas9 RNA was assessed by sequencing gDNA of eight injected embryos 4-5 dpf for locus specific alterations. Primer sequences are shown in Supplementary Table 1.

#### Zebrafish genotyping

Extraction of genomic DNA from fin clips or from whole 4-5 dpf embryos was performed by Proteinase K digestion as previously described (Westerfield 2000). 2 µl of eluted genomic DNA (approx. 50 ng/µl) was used for PCR amplification, with subsequent clean-up and Sanger sequencing. Sequences of primers used in this study are given in Supplementary Table 1. Sequencing results were analyzed with “ApE” (http://biologylabs.utah.edu/jorgensen/wayned/ape/) and CodonCode Aligner (CodonCode Corporation, Centerville, MA, USA) software packages.

#### Scanning electron microscopy

Zebrafish larvae at 5 dpf were fixed overnight at 4°C with 6.5% glutaraldehyde solution in Sörens buffer. After fixation, embryos were washed 3x in PBS and dehydrated by an uprising dilution series into acetone. Subsequently, embryos underwent critical point drying and were gold/palladium coated. Images were taken with a JEOL JSM-7500F (Akishima, Tokyo, Japan), using either LEI or SEI detectors at 10,000x magnification.

#### DASPEI staining and neuromast quantification in zebrafish larvae

For hair cells and neuromast visualization in living larvae, the fluorescent dye 2-[4-(dimethylamino)styryl]-N-ethylpyridinium iodide (DASPEI; Life Technolgies, Carlsbad, CA, USA) was used. Larvae at the desired stage (age before 120 hpf) were incubated for 15 minutes in a 0.13 mM DASPEI solution in 30% Danieau’s medium. The larvae were subsequently washed three times for one minute in 30% Danieau’s. Finally, a Tricain incubation was performed prior to imaging. The fish were mounted into a 2.5% methylcellulose/30% Danieau’s medium solution and laterally imaged under a fluorescence stereomicroscope using light-field, gfp and rfp filters. Larvae of *frmpd4*^sa12377^ strain crossings were of unknown genotype during imaging and were subsequently fixed for postmortem genotype determination.

For the quantification of the number and distance between the neuromasts of the posterior lateral line (PLL, Clusters P1-P9) in images of DASPEI stained zebrafish Fiji software was used. Distances between clusters were measured in pixels. The resulting values of the pixel distances between the neuromasts were scaled to the overall length between P1 and P9 using Microsoft excel and thus representing the distances as percent values. The calculated relative distances were statistically analyzed using OriginPro 2021 and illustrated via boxplot diagrams including data points. For standard deviation, the coefficient value was set to 1.5 and is indicated by the whiskers within the blot. Further values depicted in the box blots are the median (parallel line), the mean value (small box), and the upper and lower quartile (large box). For statistical analysis, the obtained data values were first tested for normal distribution, and the significance was determined using the Mann-Whitney-U test. The significance values were given as U values with a significance level set to be smaller than 5 % (U < 0.05). Values with a statistical significance of U ≤ 0.05 were marked with *, while a significance level of U ≤ 0.0001 is marked as ** and *** for U ≤ 0.00001.

For quantification of neuromast numbers in the laterally visible otic vesicle (OV) of the same embryos, DASPEI stained neuromast cells within the organ were manually counted subsequently. Quantification of preoptic/supraorbital neuromast clusters (ventral to the eye; indicated in control embryo Figure 3A) was performed as a control for normal cranial neuromast development. Statistical analyses were performed like PLL measurements using the Mann-Whitney-U test.

#### Immunofluorescence in zebrafish experiments

Immunofluorescence staining of axons and kinocilia in hair cells was performed according to previously established protocols (Raible and Kruse 2000; Tanimoto et al. 2011) using an acetylated Tubulin antibody (primary monoclonal mouse antibody IgG2b 6-11B-1; sc-23950, Santa Cruz Biotechnology, Antibody Registry nr. AB_628409; 1:1000 diluted in 1% sheep serum/PBST) and anti-mouse IgG Alexa 594 secondary antibody (A-1062; Thermo Fisher Scientific, Waltham, MA, USA; Antibody Registry nr. AB_2534109; 1:5000 diluted in PBST) or anti-mouse IgG Alexa Fluor 488 (A-11001; Thermo Fisher Scientific, Waltham, MA, USA; Antibody Registry nr. AB_2534069, 1:5000 diluted in PBST). For counterstaining of nuclei Hoechst 33342 (dilution 1:10000; Invitrogen/Thermo Fisher Scientific, Waltham, MA, USA) was added to the to the secondary antibody solution. F-Actin was visualized with Phalloidin-488 (1:20 dilution in PBST; Acti-stain 488, Biozol, Eching, Germany). Images were taken with a Nikon confocal laser scanning microscope A1+ and analyzed with Fiji/ImageJ software (https://fiji.sc/).

#### Confocal microscopy and imaging processing

High-resolution immunofluorescence images were taken with a Nikon confocal laser-scanning microscope A1+ setup with a D-eclipse C1si detector and corresponding SP fluorescence filters (Nikon) used for 405nm, 488nm, 561nm excitation wavelength lasers (Nikon Europe B.V.; Amstelveen, The Netherlands). For visualization of cranial or trunk region of zebrafish embryos, head and trunk were manually separated and individually mounted for confocal imaging. Standard confocal imaging utilized a Plan Apo VC 20x DIC N2 objective and recordings at 2048x2048 x/y-pixel size. Cranial region images included up to 55 z-stack images (overall imaging depth ∼110µm), while trunk region images included up to 30 z-stack images (overall imaging depth ∼60µm). Voxel size was set to 0.16x0.16x2.5µm up to 0.16x0.16x5µm. Acquired.nd2 files from NIS-Elements software (Nikon) were further processed with imageJ/Fiji. For visualization of neuromast centers in the otic vesicle or in the lateral line maximum intensity projections of ROIs were calculated. “Despeckle” routine was used to reduce image noise of confocal stack images. Color overlay images were processed for optimal orientation, brightness, contrast and were colored-coded by different LUTs to get best visualization.

#### Zebrafish startle response test

Startle response tests have been conducted as previously reported (Bhandiwad et al. 2013; Wang et al. 2015). In short, single adult zebrafish (aged between 5 and 7 months, mixed sex ratios) were investigated in an experimental set up and response to a given sound stimulus (frequency: 4.4 kHz; duration: 20ms; sound-intensity levels in air: 0.01W/m^2^ (100dB) to 0.1W/m^2^ (>110dB): 4 to 6 sound pulses in a 2 minute time frame) was digitally recorded after habituation with a high speed camera at 50 frames/second. Release of a startle response within a timeframe of 200 ms (10 frames) after the sound stimulus and reaction time was quantified (Software: VLC media player, Adobe Premier Elements 14 and ImageJ/Fiji). Reaction time was quantified by measuring frames between sound stimulus detection by the sound meter and animal reaction. Two independent experiments were conducted and overall, 12 fish per genetic group (wild type (WT); *frmpd4^sa12377/+^*; *frmpd4^sa12377/sa12737^*) were investigated.

#### *In vivo* RNA overexpression

RNA overexpression in zebrafish embryos was performed by microinjections of human *FRMPD4* capped mRNA (final concentration 25 ng/µl in injection solution) into one-cell stage zebrafish embryos. Injection controls were performed, by substituting mRNA with water. To produce WT and patient-specific *FRMPD4* capped mRNA, full-length *FRMPD4* CDS (Dharmacon Clone-ID 8322752; Accession nr.: BC113700; Horizon Discovery) was subcloned into pCS2p+ (pCS2P+ was a gift from Marc Kirschner; Addgene plasmid #17095; http://n2t.net/addgene:17095; RRID:Addgene 17095). Patient-specific variants were introduced via site-directed mutagenesis (NEB Q5® Site-Directed Mutagenesis Kit; New England Biolabs, Ipswich, MA, USA) and validated via Sanger-Sequencing. Capped mRNA was produced via the mMESSAGE mMACHINE™ SP6 Transcription kit (Thermo Fisher Scientific, Waltham, MA, USA), purified and validated by RNA gel electrophoresis.

### Mouse methods

#### RT-qPCR analysis of *Frmpd4* expression in mouse tissues

RNA was extracted from multiple tissues from WT male C57BL/6 (P7) mice following standard protocols (Vikhe Patil et al. 2015). RNA quality was assessed with a NanoDrop spectrophotometer (NanoDrop Technologies, Wilmington, DE). cDNA was produced using the SuperScript III First-Strand Synthesis SuperMix RT-PCR kit (Invitrogen, Karlsruhe, Germany). cDNA from different mouse tissues were generated (1 µg whole organ RNA) and analyzed by qPCR. The *Frmpd4* (NM_001033330) and *Hprt* cDNA regions of interest were by amplified using standard PCR conditions using primers mm *Frmpd4* Ex10-11 F and R and mm *Frmpd4* Ex16-17 F and R. RT-qPCR of *Frmpd4* and the endogenously expressed genes *Eef2* (NM_007909) and *Tbp* (NM_013684) were performed according to standard protocols. Primers are listed in Supplementary Table 1.

Reverse transcription was performed using 1 μg RNA per sample and FIREScript RT cDNA synthesis kit (Solis BioDyne, Tartu, Estonia) according to manufacturer’s instructions. Relative expression levels were generated using qPCR HOT Fire Pol Eva Green qPCR Mix Plus (Solis BioDyne, Tartu, Estonia) according to standard protocols. Two different intron-spanning primer sets were selected for analysis of *Frmpd4* expression (targeting exons 8-9, 10-11 and 16-17) and *Eef2*, as well as *Tbp* were used for cDNA normalization. The PCR reactions were performed in technical triplicates. The qPCR reactions were performed using 364-well-plates and the Quantstudio Real-Time PCR system (Life Technologies, Darmstadt, Germany). The following program was used: 95°C 15 min, 40x [95°C, 15 s, 60°C 20 s, 72°C 20 s] followed by the generation of a melting curve using temperatures from 60°C until 95°C. Primer sequences are listed in Supplementary Table 1.

#### *In silico* analysis of *Frmpd4* expression in the cochlea and spiral ganglion neurons (SGNs) in the WT mouse

We performed an *in silico* analysis of the expression of *Frmpd4* during mouse development using various publicly available RNA-seq datasets. First, we studied the expression of *Frmpd4* across several cochlear tissues in the WT P8 mouse using the dataset “Xenium Analysis of P8 WT Mouse Inner Ear Sections (Violin),” generated using the Xenium analyzer (10x Genomics). This dataset was generated by fixing P8 WT cochleae that were harvested, fixed in paraformaldehyde, decalcified, and embedded in paraffin wax. Tissue was sectioned in 5 µm sections and placed on Xenium slides. The standard Xenium protocol was followed and custom probes were applied. A Xenium Analyzer was used to decode the probes. We next studied SGN expression, using the Gene Expression Omnibus (GEO) Series GSE132925 (Li et al. 2020) that assessed expression in WT mice at embryonic day (E)15.5, P1, P8, P14, and P30. This dataset was generated using RNA-seq on manually dissected WT mouse SGNs at these five distinct ages. In order to identify SGN-specific genes, the transcriptomes of the SGNs were compared with those of the inner ear hair cells at P12 and glial cells at P8. Finally, analysis of *Frmpd4* expression in SGN types Ia, Ib, Ic, and 2 were analyzed using GEO Series GSE114997 (Shrestha et al. 2018). This previously generated dataset isolated cochlear sensory neurons from P25 to P27 mice (Genotypes: *Bhlhb5^Cre^/+*; *Ai14/+* and *Bhlhb5^Cre^/+*; *Ai14/+*; *Vglut3^-/-^*). Gene expression was measured using scRNA-seq. The dataset consists of gene expression from 63 type Ia, 71 type Ib, 45 type Ic and 7 type II neurons. All data were visualized in the gene expression analysis resource (gEAR) portal (Orvis et al. 2021).

#### Immunofluorescence in mouse experiments

For immunohistochemical analysis of cochlea, mice were deeply anesthetized with CO_2_ and perfused intracardially with 4% PFA (Roth, #03353) in 1 M PBS pH 7.4 at room temperature. Cochleae were postfixed over night with 4% PFA in 1 M PBS pH 7.4 and afterwards decalcified in 125 mM EDTA (Invitrogen, #AM9262) for 48 h, while EDTA was changed after 24h. For whole mount staining, the cochlear turns were separated and covered in a 24 well plate with 1% PFA in 1 M PBS till preparations are finished. For cryosections, the mouse cochleae were incubated in ascending concentrations of sucrose (Roth, #90971) along with an additional suspension of Compound Tissue Tek (Sakura, #4583). The cochleae were afterwards embedded and frozen at -20°C in pure Compound Tissue Tek and cut in 5 μm sections. The organ of Corti and the spiral ganglia of whole mount and cryosections was first blocked and permeabilized with a solution of 10% normal horse serum (Merck, #H0146), 1% bovine serum albumin (Roth, #80762), 1% Triton X-100 (Serva, #37240), and 0.1% Tween20 (Sigma, #P1379) in 1 M PBS pH 7.4 at room temperature. Afterwards, the tissue was incubated with a primary antibody solution containing 3% normal horse serum, 1% bovine serum albumin, 0.3% Triton X-100, and 0.1% Tween20 with primary antibodies for βII-tubulin (mouse monoclonal, R&D System, #MAB1192 clone Tuj1, dilution 1:1000) and FRMPD4 (rabbit, polyclonal, dilution 1:500) (Hu et al. 2012) incubated overnight at 4°C. On the next day, primary antibodies were detected with secondary antibody Alexa488 (donkey anti-mouse IgG (H+L) Highly Cross-Adsorbed Secondary Antibody, Invitrogen, A21206), and Alexa-555 (donkey anti-rabbit IgG (H+L) Highly Cross-Adsorbed Secondary Antibody, Invitrogen, A32794 or goat anti-rabbit IgG (H+L) Cross-Adsorbed Secondary Antibody, Invitrogen, A21428; dilution 1:1000) and DAPI (dilution 1:5000; Invitrogen). Counterstaining of F-Actin was performed by fluorescent conjugated Phalloidin staining according to manufacturer instructions. Images of stained cryosections were taken with a Nikon confocal laser scanning microscope A1+ and analyzed with Fiji/ImageJ software. Images of the whole mount staining were taken with an Olympus IX81 microscope equipped with an Olympus FV1000 confocal laser scanning system, an FV1000 SPD spectral detector and diode lasers operating at 473, 559 and 651 nm. Images were acquired using an Olympus UPLSAPO 40x objective (oil, numerical aperture: 1.3). For high-resolution confocal scanning, a pinhole aperture representing a diffraction disk was used. Whole-mount images of the organ of Corti were taken at 300 nm intervals along the z-axis. The z-stacks, brightness and contrast of the images were adjusted using ImageJ for better visualization.

#### Auditory brainstem response (ABR) testing in mice

Hearing thresholds were measured by click and tone burst ABR (6, 8, 12, 16, and 24 kHz) for *Frmpd4*^-/-^ and WT mice. ABR testing from 14 male mice included four and two *Frmpd4*^-/-^ mice and six and two WT mice at 3 and 5 months of age, respectively. ABRs were measured as previously described (Lauer and May 2011; McGuire et al. 2015; Schrode et al. 2022; Schrode et al. 2018). Mice were anesthetized with 100 mg/kg ketamine and 10 mg/kg xylazine and placed on a heating pad to maintain a temperature of 37°C. Reference, active, and ground platinum needle electrodes were placed subcutaneously behind the left pinna, at the dorsal midline of the skull, and in the left hind leg, respectively. Responses were amplified and filtered from 300 to 3000 Hz and averaged over 300 repetitions. Stimulus generation and ABR measurements were controlled and collected using computer programming modules (Tucker-Davis Technologies, Alachua, FL, USA) and custom Matlab-based software. Averaged ABR waveforms were obtained by presenting clicks or 5 ms pure-tone pips (0.5 ms cos^2^ onset/offset) at a rate of 20 stimuli/s presented in free field at a distance of 30 cm from the vertex of the skull. Hearing thresholds (dB SPL) were measured by presenting a descending series of stimulus levels in 5- or 10-dB steps until no discernable response was detected. Maximum ABR thresholds were capped at 108 dB SPL due to system output limits. The ABR signal magnitude was calculated from averaged peak-to-peak voltage in a 10-ms time window beginning 1 ms after stimulus onset. Threshold was determined to be the sound level at which the ABR amplitude exceeded the background noise by two standard deviations. A mixed model was used to test for significant differences in ABR thresholds between *Frmpd4*^-/-^ and WT mice, including subject ID as a random effect to account for repeated measures.

### Supplementary Tables

**Supplementary Table 1.**
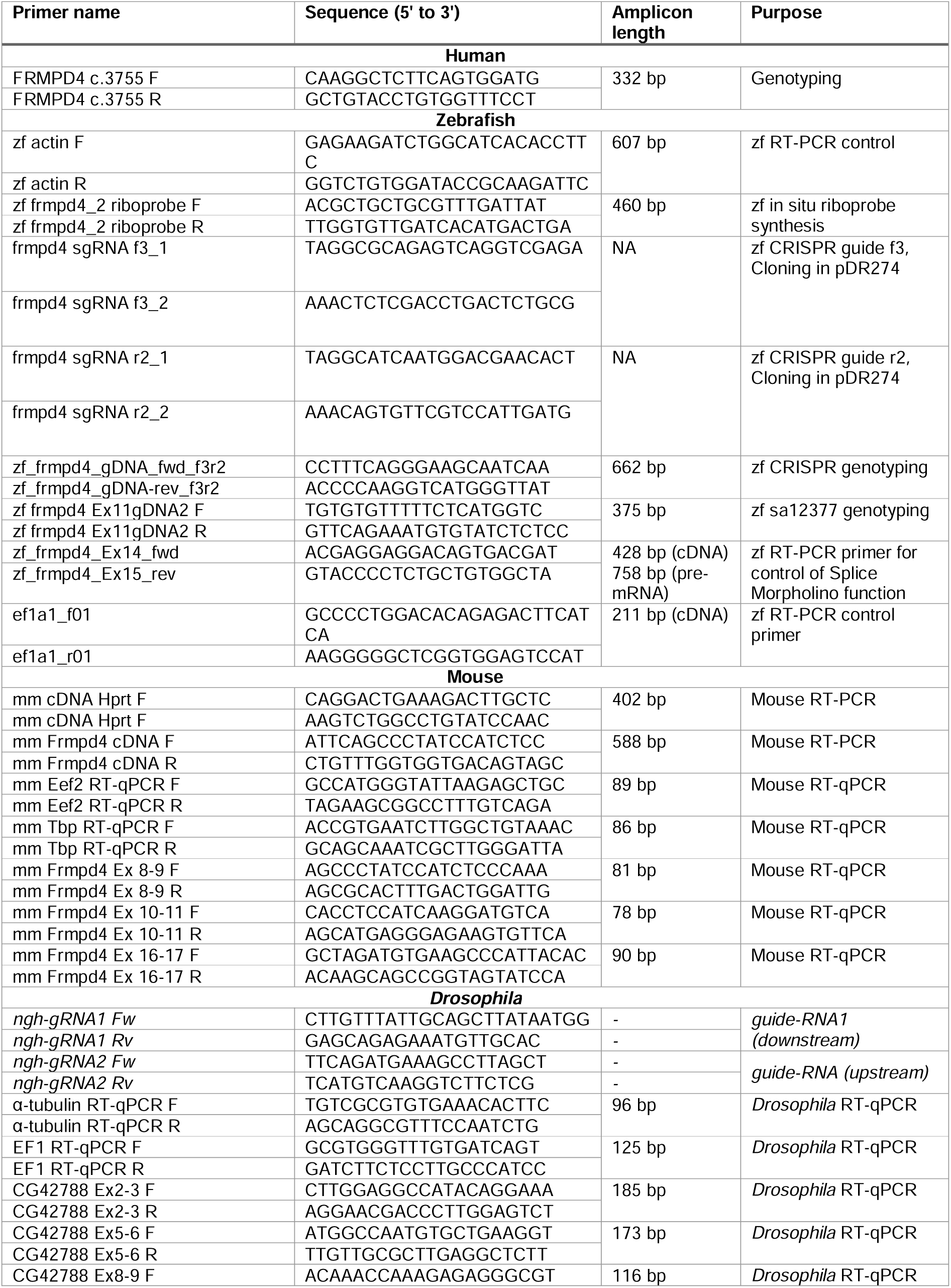
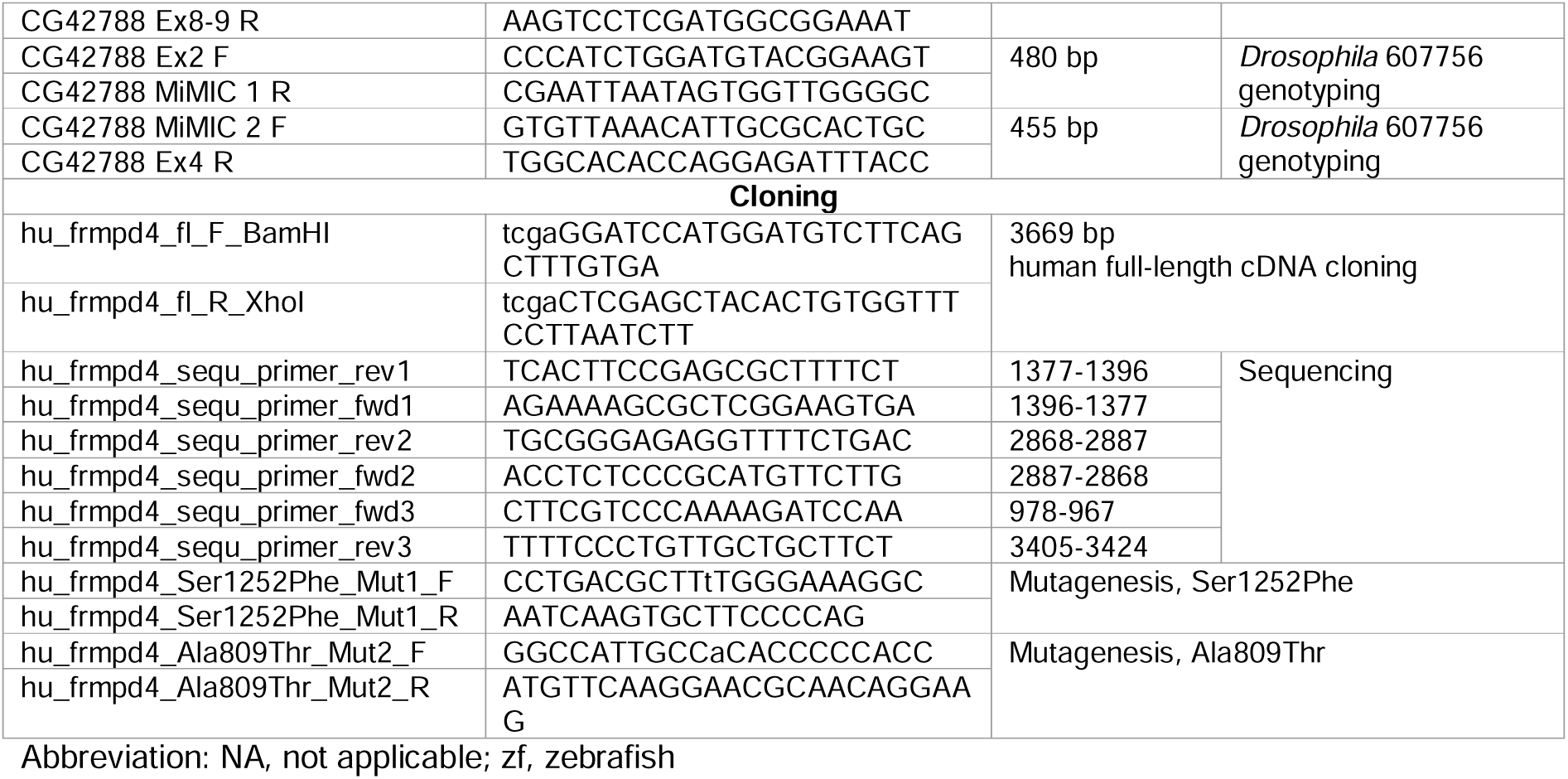
Primers used in the study.

**Supplementary Table 2.** Exome filtering details for Family 1.

|  |  |
| --- | --- |
| Total combined familial transcript variants | 304,240 |
| Exonic transcript variants | 11,846 |
| Transcript variants with Qscore $\geq 30$ | 9,508 |
| Transcript variants with a MAF $\leq 0.01$ (1000 Genomes Project and Exome Variant Server) | 4,703 |
| In house MAF $\leq 0.01$ (n = 511 exomes) | 1,819 |
| Exclude transcript variants in artifact-prone genes* | 1,758 |
| Total missense, stop gain, stop loss, start gain, start loss, splice, indel, total count transcript variants | 1,327 |
| Total synonymous transcript variants | 401 |
| Transcript-independent variants: Missense, stop gain, stop loss, start gain, start loss, splice, indel, total count | 714 |
| Pedigree-filtering variant analysis |  |
| Autosomal recessive mode of inheritance: |  |
| Homozygous in III:2 and III:3 | 0 |
| Heterozygous in II:2 and II:3 and not inherited from a parent | 0 |
| Compound heterozygous in III:2 and III:3 | 0 |
| Autosomal dominant mode of inheritance: |  |
| Heterozygous in III:2, III:3, II:3, absent in III:2, coverage $\geq 20x$, MAF $\leq 0.00002^{a,b}$ | 14 |
| Heterozygous/Hemizygous variants predicted pathogenic by 2/3 of *in silico* tools | 3 |

\*HLAs, MAGEs, MUCs, NBPFs, ORs, PRAMEs
<sup>a</sup>Adhering to guidelines for variant filtering from Oza et al., 2018.
<sup>b</sup>Shown in Supplementary Table 3.

**Supplementary Table 3.**
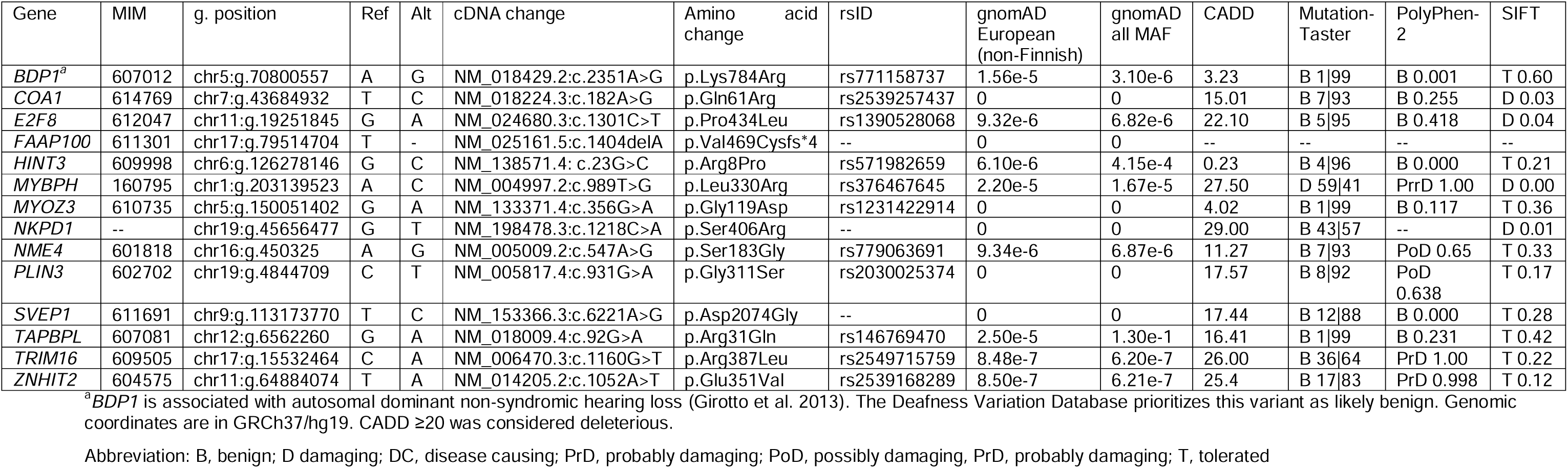
Heterozygous variants identified in the affected individuals in Family 1 that were inherited from a healthy parent.

| Gene | MIM | g. position | Ref | Alt | cDNA change | Amino acid change | rsID | gnomAD European (non-Finnish) | gnomAD all MAF | CADD | Mutation-Taster | PolyPhen-2 | SIFT |
| --- | --- | --- | --- | --- | --- | --- | --- | --- | --- | --- | --- | --- | --- |
| *BDP1<sup>a</sup>* | 607012 | chr5:g.70800557 | A | G | NM_018429.2:c.2351A>G | p.Lys784Arg | rs771158737 | 1.56e-5 | 3.10e-6 | 3.23 | B 1\|99 | B 0.001 | T 0.60 |
| *COA1* | 614769 | chr7:g.43684932 | T | C | NM_018224.3:c.182A>G | p.Gln61Arg | rs2539257437 | 0 | 0 | 15.01 | B 7\|93 | B 0.255 | D 0.03 |
| *E2F8* | 612047 | chr11:g.19251845 | G | A | NM_024680.3:c.1301C>T | p.Pro434Leu | rs1390528068 | 9.32e-6 | 6.82e-6 | 22.10 | B 5\|95 | B 0.418 | D 0.04 |
| *FAAP100* | 611301 | chr17:g.79514704 | T | - | NM_025161.5:c.1404delA | p.Val469Cysfs*4 | -- | 0 | 0 | -- | -- | -- | -- |
| *HINT3* | 609998 | chr6:g.126278146 | G | C | NM_138571.4: c.23G>C | p.Arg8Pro | rs571982659 | 6.10e-6 | 4.15e-4 | 0.23 | B 4\|96 | B 0.000 | T 0.21 |
| *MYBPH* | 160795 | chr1:g.203139523 | A | C | NM_004997.2:c.989T>G | p.Leu330Arg | rs376467645 | 2.20e-5 | 1.67e-5 | 27.50 | D 59\|41 | PrrD 1.00 | D 0.00 |
| *MYOZ3* | 610735 | chr5:g.150051402 | G | A | NM_133371.4:c.356G>A | p.Gly119Asp | rs1231422914 | 0 | 0 | 4.02 | B 1\|99 | B 0.117 | T 0.36 |
| *NKPD1* | -- | chr19:g.45656477 | G | T | NM_198478.3:c.1218C>A | p.Ser406Arg | -- | 0 | 0 | 29.00 | B 43\|57 | -- | D 0.01 |
| *NME4* | 601818 | chr16:g.450325 | A | G | NM_005009.2:c.547A>G | p.Ser183Gly | rs779063691 | 9.34e-6 | 6.87e-6 | 11.27 | B 7\|93 | PoD 0.65 | T 0.33 |
| *PLIN3* | 602702 | chr19:g.4844709 | C | T | NM_005817.4:c.931G>A | p.Gly311Ser | rs2030025374 | 0 | 0 | 17.57 | B 8\|92 | PoD 0.638 | T 0.17 |
| *SVEP1* | 611691 | chr9:g.113173770 | T | C | NM_153366.3:c.6221A>G | p.Asp2074Gly | -- | 0 | 0 | 17.44 | B 12\|88 | B 0.000 | T 0.28 |
| *TAPBPL* | 607081 | chr12:g.6562260 | G | A | NM_018009.4:c.92G>A | p.Arg31Gln | rs146769470 | 2.50e-5 | 1.30e-1 | 16.41 | B 1\|99 | B 0.231 | T 0.42 |
| *TRIM16* | 609505 | chr17:g.15532464 | C | A | NM_006470.3:c.1160G>T | p.Arg387Leu | rs2549715759 | 8.48e-7 | 6.20e-7 | 26.00 | B 36\|64 | PrD 1.00 | T 0.22 |
| *ZNHIT2* | 604575 | chr11:g.64884074 | T | A | NM_014205.2:c.1052A>T | p.Glu351Val | rs2539168289 | 8.50e-7 | 6.21e-7 | 25.4 | B 17\|83 | PrD 0.998 | T 0.12 |

<sup>a</sup>*BDP1* is associated with autosomal dominant non-syndromic hearing loss (Giotto et al. 2013). The Deafness Variation Database prioritizes this variant as likely benign. Genomic coordinates are in GRCh37/hg19. CADD $\geq 20$ was considered deleterious.
Abbreviation: B, benign; D damaging; DC, disease causing; PrD, probably damaging; PoD, possibly damaging; PrD, probably damaging; T, tolerated

**Supplementary Table 4.** Variants in known hearing loss genes and candidate genes identified in affected individuals through exome sequencing.

| Family 1 pedigree filtering |  |  |  |  |  |  |  |  |  |  |  |  |  |
| --- | --- | --- | --- | --- | --- | --- | --- | --- | --- | --- | --- | --- | --- |
| Chr | bp (hg19) | Ref | Alt | Zyg | Gene | cDNA change | aa change | rs | gnomAD MAF | CADD | MT | PolyPhen-2 | SIFT |
| 11 | 17,531,325 | G | A | Het | *USH1C* | NM_153676.3:c.1591C>T | p.Arg531Cys | rs140528164 | 0.00007379 | 28.3 | DC | PD | T |
| X | 12,736,700 | C | T | Hemi | *FRMPD4* | NM_014728.3:c.3755C>T | p.Ser1252Phe | NR | NR | 25.4 | DC | PD | D |
| X | 18,972,407 | C | T | Hemi | *PHKA2* | NM_000292.2:c.202G>A | p.Asp68Asn | rs866514095 | 0.00002495 | 25.1 | DC | PD | D |
| Family 2, individual IV:1 |  |  |  |  |  |  |  |  |  |  |  |  |  |
| 1 | 110,019,548 | C | CTA | Hom | *SYPL2* | NM_001040709.1:c.406_407dup | p.Leu137fsThrfs*21 | NR | NR | NR | DC | NR | NR |
| 10 | 73,570,263 | C | T | Het | *CDH23* | NM_022124.5:c.9014C>T | p.Ala3005Val | rs188966938 | 0.00001605 | 22.5 | DC | B | D |
| 12 | 112,326,421 | G | A | Hom | *MAPKAPK5* | NM_139078.2:c.1099G>A | p.Gly367Ser | NR | NR | 34 | DC | DC | D |
| 17 | 76,046,996 | G | T | Hom | *TNRC6C* | NM_001142640.2:c.2483G>T | p.Gly828Val | NR | NR | 23.6 | DC | NR | D |
| X | 127,350,003 | G | A | Hemi | *FRMPD4* | NM_014728.3:c.2425G>A | p.Ala809Thr | NR | 0.000005600 | 27.9 | DC | PD | T |

Abbreviations: aa, amino acid; alt, alternate base; bp, base pair position (GRCh37, hg19); CADD, Combined Annotation Dependent Depletion; Chr, chromosome; D, deleterious; DC, disease causing; gnomAD MAF, genome aggregation database minor allele frequency; NR, not reported; ref, reference; T, tolerated; zyg, zygosity

**Supplementary Table 5.** Exome filtering details for Family 2, individual IV:1.

|  |  |
| --- | --- |
| Total variants | 99,819 |
| Exonic transcript variants | 25,845 |
| Transcript variants with Qscore $\geq 30$ | 15,043 |
| Transcript variants with a MAF $\leq 0.00007$ (1000 Genomes Project and Exome Variant Server) | 869 |
| Exclude transcript variants in artifact-prone genes* | 507 |
| Total missense, stop gain, stop loss, start gain, start loss, splice, indel | 348 |
| Total synonymous variants | 159 |
| Autosomal recessive mode of inheritance predicted to be pathogenic by 2/3 of *in silico* tools: |  |
| Homozygous variants | 4 |
| Compound heterozygous variants | 0 |
| X-linked mode of inheritance pathogenic by 2/3 of *in silico* tools: |  |
| Hemizygous variants | 1 |

**Supplementary Table 6.** Homozygous variants identified in individual IV:1 from Family 2.

| Gene | MIM | g. position | Ref | Alt | cDNA change | Amino acid change | rsID | gnomAD all ME | gnomAD all MAF | CADD score, scaled | Mutation-Taster | PolyPhen-2 | SIFT |
| --- | --- | --- | --- | --- | --- | --- | --- | --- | --- | --- | --- | --- | --- |
| *DPP4* | 102720 | chr2:g.162,903,933 | T | C | NM_001935.3:c.173A>G | p.Tyr58Cys | rs756475707 | 0 | 2.54e-6 | 23.60 | B 14\|86 | PrD 1.0 | T 0.13 |
| *MAPKAPK5* | 606723 | chr12:g.112,326,421 | G | A | NM_139078.2:c.1099G>A | p.Gly367Ser | -- | 0 | 0 | 23.90 | D 58\|42 | PrD 1.0 | D 0.00 |
| *PRDM9* | 609760 | chr5:g.23,522,762 | C | A | NM_020227.2:c.650C>A | p.Ala217Asp | -- | 0 | 0 | 22.00 | B 13\|87 | PrD 0.983 | T 0.08 |
| *SYPL2* | -- | chr1: g.110,019,548 | C | CTA | NM_001040709.1:<br>c.406_407dupTA | p.Leu137Thrfs<br>*21 | -- | -- | -- | -- | -- | -- | -- |
| *TNRC6C* | 610741 | chr17:g.76,046,996 | G | T | NM_001142640.1:c.1853G<br>>T | p.Gly618Val | -- | -- | -- | 23.60 | B 21\|79 | -- | D 0.00 |

Abbreviations: B, benign; D damaging; DC, disease causing; ME, Middle Eastern; PrD, probably damaging; PoD, possibly damaging, PrD, probably damaging; T, tolerated

**Supplementary Table 7.** FRMPD4 variants with clinical summary.

| ***FRMPD4* variant** | **Inheritance** | **Phenotype** | **Reference** |
| --- | --- | --- | --- |
| ~632 kb duplication including part of *FRMPD4* (Exon 1 duplicated) | Maternal | Mild ID and autism | (Honda et al. 2010) |
| ~66 kb microdeletion including exon 2 | Maternal | Delayed gross motor development, delayed speech and language development, severe to profound ID, autism, hypotonia, spasticity, brisk deep tendon reflexes, ataxia, reduced occipitofrontal circumference, epilepsy, strabismus | (Piard et al. 2018) |
| c.380C>T, p.Pro127Leu (Exon 4) | Maternal | ID | (Trujillano et al. 2017) |
| c.856C>T, p.Arg286* (Exon 9) | Maternal | Delayed gross motor development, delayed speech and language development, moderate ID, hyperactivity (aggressiveness/autism) | (Piard et al. 2018) |
| c.1657T>C, p.Cys553Arg (Exon 15)<sup>1</sup> | *de novo* | Mild to moderate ID, developmental delay, mild facial features, absent speech, autism spectrum disorder | (Hu et al. 2016) |
| c.1657T>C, p.Cys553Arg (Exon 15)<sup>1</sup> | *de novo* | Delayed gross motor development, delayed speech and language development, severe ID, autism, strabismus | (Piard et al. 2018) |
| c.1772A>C, (p.Glu591Ala) (Exon 15) | Maternal | Severe ID and language impairment | (Pan et al. 2024) |
| c.1852delT, p.Cys618Valfs*8 (Exon 15) | Maternal | Mild to severe ID with variable seizures, lack of speech/poor speech, behavioural problems | (Hu et al. 2016) |
| c.2425G>A, p.Ala809Thr (Exon 15) | Maternal | Non-syndromic sensorineural hearing loss | Present study |
| c.2641G>A, p.Val881Met (Exon 15) | Maternal | Isolated epilepsy | (Li et al. 2024) |
| c.2800C>T, p.Arg934Cys (Exon 16) | Maternal | Isolated epilepsy | (Li et al. 2024) |
| c.2816C>T, p.Thr939Ile (Exon 16) | Maternal | Epilepsy with ID | (Li et al. 2024) |
| c.3061T>C, p.Tyr1021His (Exon 16) | Maternal | Isolated epilepsy | (Li et al. 2024) |
| c.3623T>C, p.Phe1208Ser (Exon 16) | Maternal | Isolated epilepsy | (Li et al. 2024) |
| c.3755C>T, p.Ser1252Phe (Exon 16) | Maternal | Non-syndromic sensorineural hearing loss | Present study |

*FRMPD4* (NM\_001368397.1) has 17 exons; <sup>1</sup> Same *de novo* variant seen in two patients
Abbreviation: ID, intellectual disability

### Supplementary Figures

**Supplementary Figure 1.**
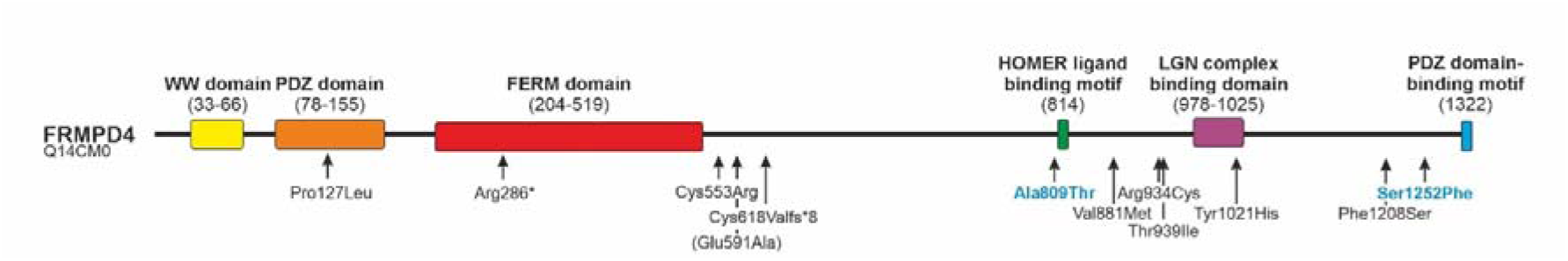
Schematic overview of *FRMPD4* protein domain structure and disiease associated genetic variants reported to date Variants exclude (Honda et al. 2010; Piard et al. 2018). Numbers in brackets indicate amino acid positions in human protein (UniProt ID: Q14CM0). WW domain: rsp5-domain, WWP repeating motif (PFAM: <u>PF00397</u>; InterPro: <u>IPR001202</u>); PDZ domain (PFAM: <u>PF00595</u>; InterPro: <u>IPR001478</u>), FERM domain (PFAM: <u>PF09379</u>; InterPro: <u>IPR000299</u>), Homer ligand binding motif (PPPGFRD; InterPro: <u>IPR019588</u>), LGN complex binding domain (InterPro: <u>IPR031938</u>), PDZ domain-binding motif (InterPro: <u>IPR001478</u>) (Lee et al. 2008; Takayanagi et al. 2015).

**Supplementary Figure 2.**
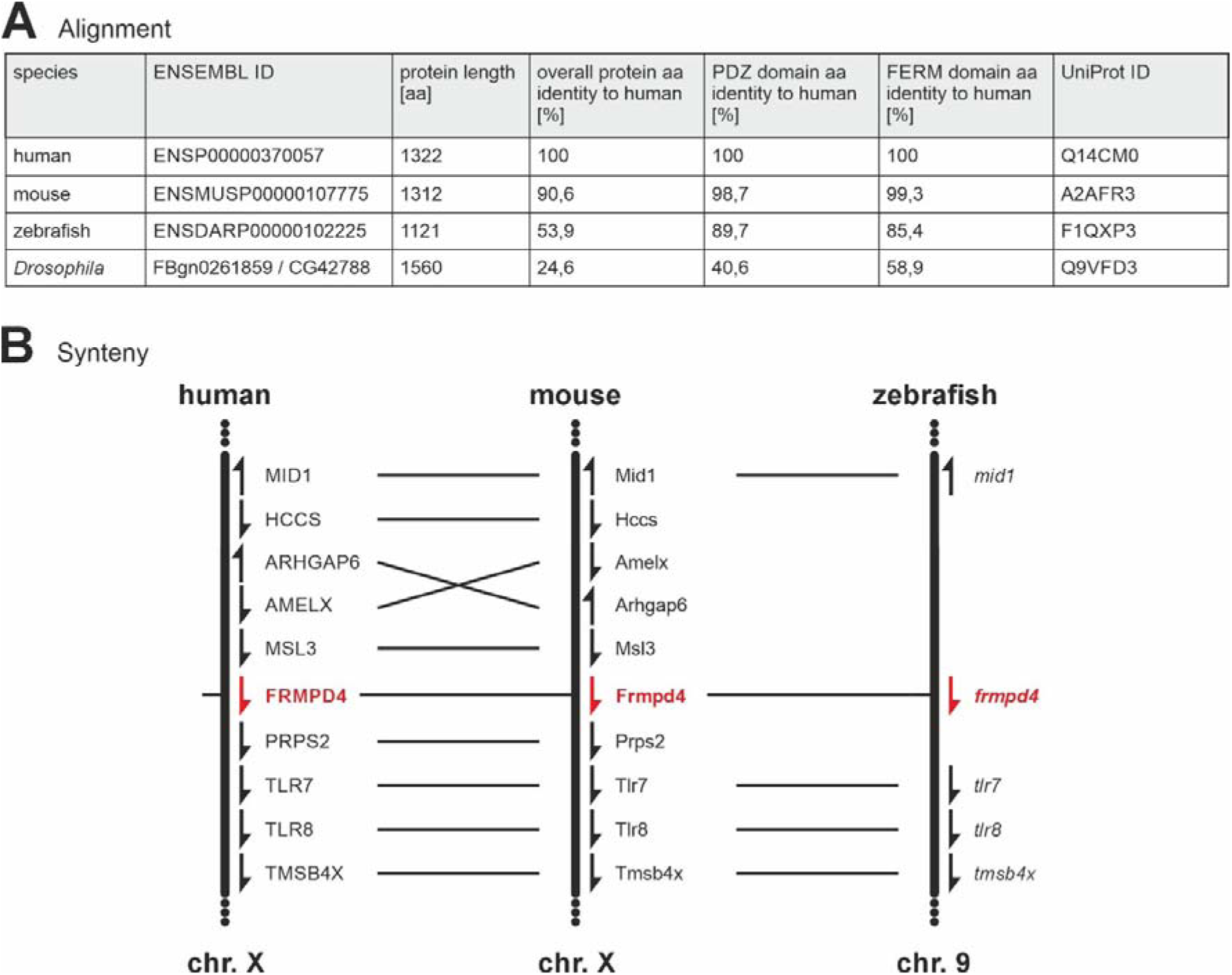
Evolutionary conservation of Frmpd4 orthologues. **(A)** Amino acid alignment of human, mouse, zebrafish and drosophila frmpd4 orthologous. **(B)** Analysis of gene locus synteny in the human, mouse and zebrafish genome.

**Supplementary Figure 3.**
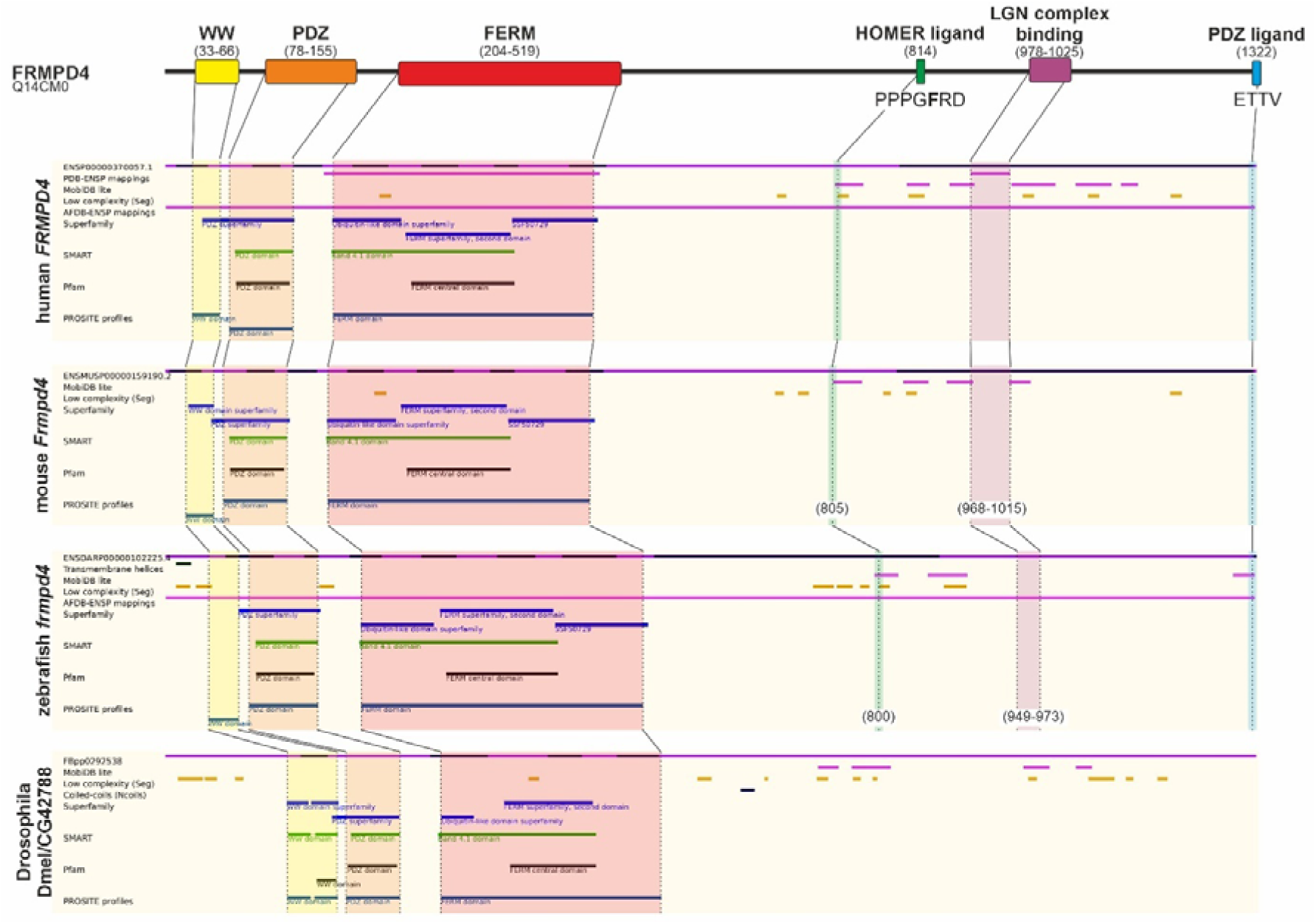
Summary of FRMPD4 protein domains and domain conservation in orthologous. Visualization of FRMPD4 protein domains. Given numbers are amino acid positions in human FRMPD4 (Uniprot ID: Q14CM0). Ensembl protein tracks and predicted domain structures of mouse, zebrafish, and *Drosophila* orthologues are aligned to human FRMPD4. Different colorations imply conserved domains.

**Supplementary Figure 4.**
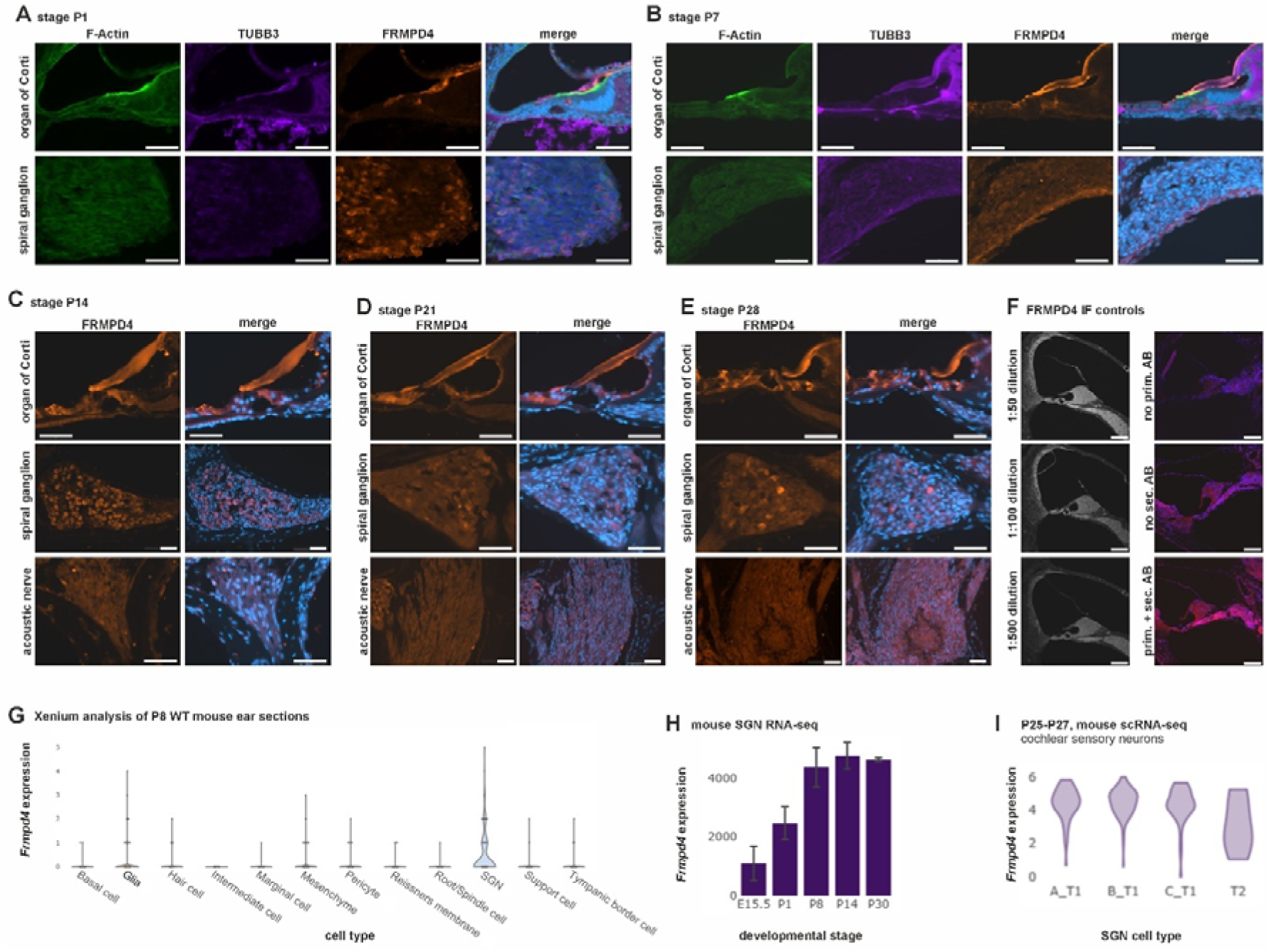
Additional mouse expression data. IF on different mouse stages and controls. Localization of FRMPD4, F-Actin and TUBB3 visualized by immunofluorescence in the organ of Corti, in the spiral ganglion and in the acoustic nerve show of mice neonates (stages **(A)**: P1; **(B)**: P7; **(C)**: P14; **(D)**: P21; **(E)**: P28). **(F)** FRMPD4 IF antibody dilution series and IF controls (no primary, no secondary, with primary and secondary antibody; similar fluorescence settings; FRMPD4 is visualized in red, nuclear counterstaining with DAPI). 5 µm sections. Scale bars = 50µm. **(G)** Violin plots of gene expression from 10x genomics xenium analysis shows highest expression in the spiral ganglion neurons (SGNs) in the P8 wild type mouse cochlea. **(H)** The expression of *Frmpd4* in the mouse SGN increases between E15.5 to P8 and then remains stable. Data were previously generated (Li et al. 2020). **(I)** Single-cell RNA sequencing shows expression of *Frmpd4* in all SGN sub-types. Data were generated previously (Shrestha et al. 2018). All data were visualized in the gene expression analysis resource (gEAR) portal (Orvis et al. 2021).

**Supplementary Figure 5.**
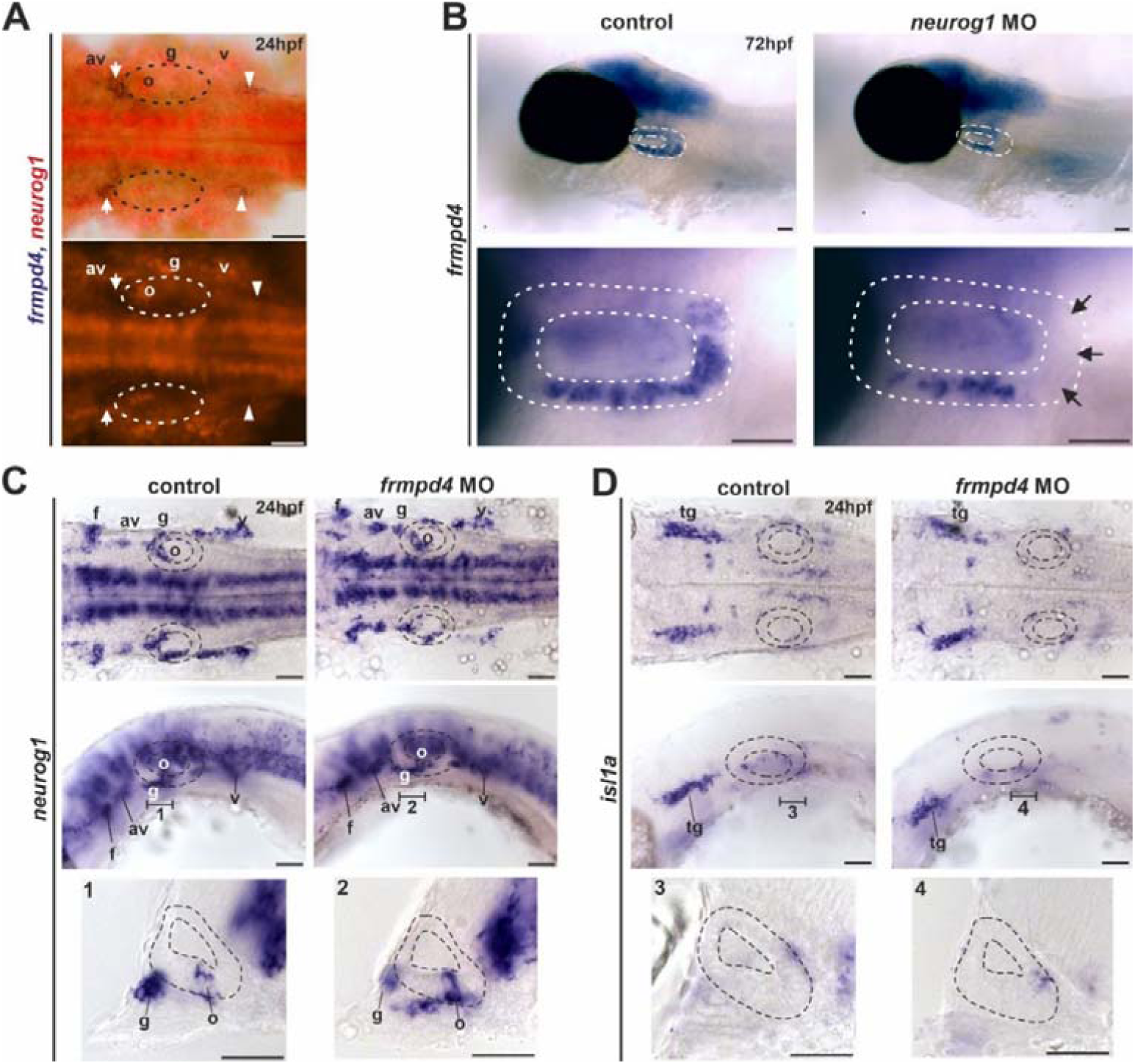
*frmpd4* expression partly localizes with *neurog1* in the otic vesicle and is regulated by *neurog1.* **(A)** Expression of *frmpd4* (stained blue) and *neurog1* (stained red) co-localize in the otic vesicle of 24hpf embryos (white arrows), but not in posterior lateral line precursors (white arrowheads). **(B)** Morpholino knockdown of *neurog1* results reduction of *frmpd4* expression exclusively in the posterior otic vesicle (black arrows; MO concentration: 0.25 mM; changed *frmpd4* expression pattern in control group: 0/12; changed *frmpd4* expression pattern in *neurog1* MO group: 12/28). Expression of *neurog1* **(C)** and *isl1* **(D)** in the otic vesicle is not influenced, by *frmpd4* knockdown (MO concentration: 0.5mM; investigated embryos per group and gene: >5). The otic vesicle is outlined with dashed lines. av - anteroventral lateral line placode; f - facial epibranchial placode; g -glossopharyngeal epibranchial placode; o - octaval/statoacoustic ganglion precursors; tg - trigeminal ganglion; v - vagal epibranchial placode. Scale bars indicate 50µm.

**Supplementary Figure 6.**
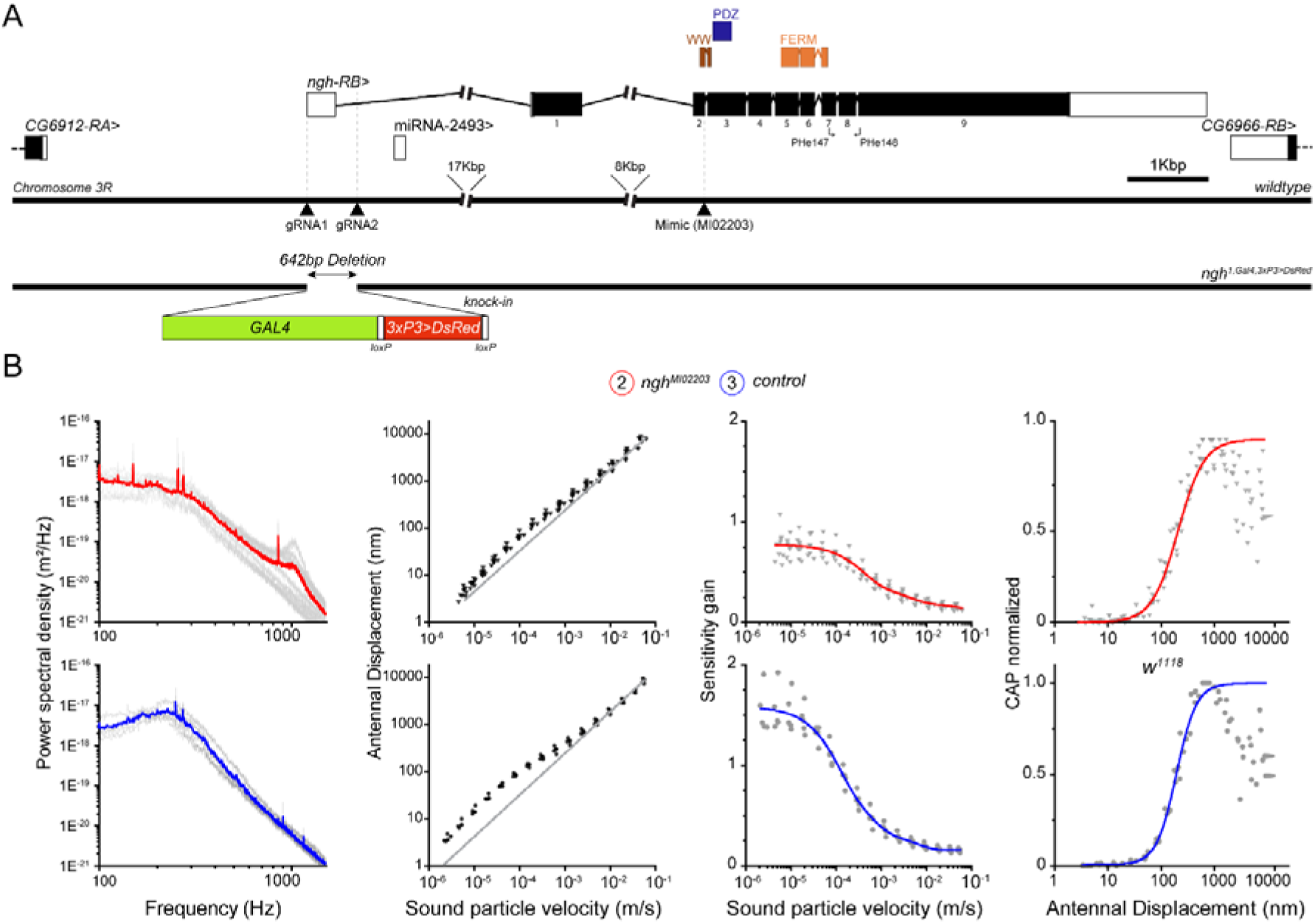
Overview of the *Drosophila* CG42788-RB gene locus and sound perception quantification. **(A)** Scheme of the genetic locus of CG42788-RB, the *FRMPD4* orthologue in *Drosophila*. Coding and non-coding exons are marked in black and white boxes; functional domains are shown with colored boxes. Gal4-3xP3-DsRed knock-in integration site and corresponding gRNA sites (spanning a 642bp deletion, including exon 1 and parts of intron 1/2) are shown at the bottom. **(B)** Quantification of Power spectral density to Frequency, Antennal Displacement to Sound particle velocity, Sensitivity gain to Sound particle velocity and CAP normalized to Antennal Displacement in *ngh (nicht gut hörend, marked in red)* and control (marked in blue) flies.

**Supplementary Figure 7.**
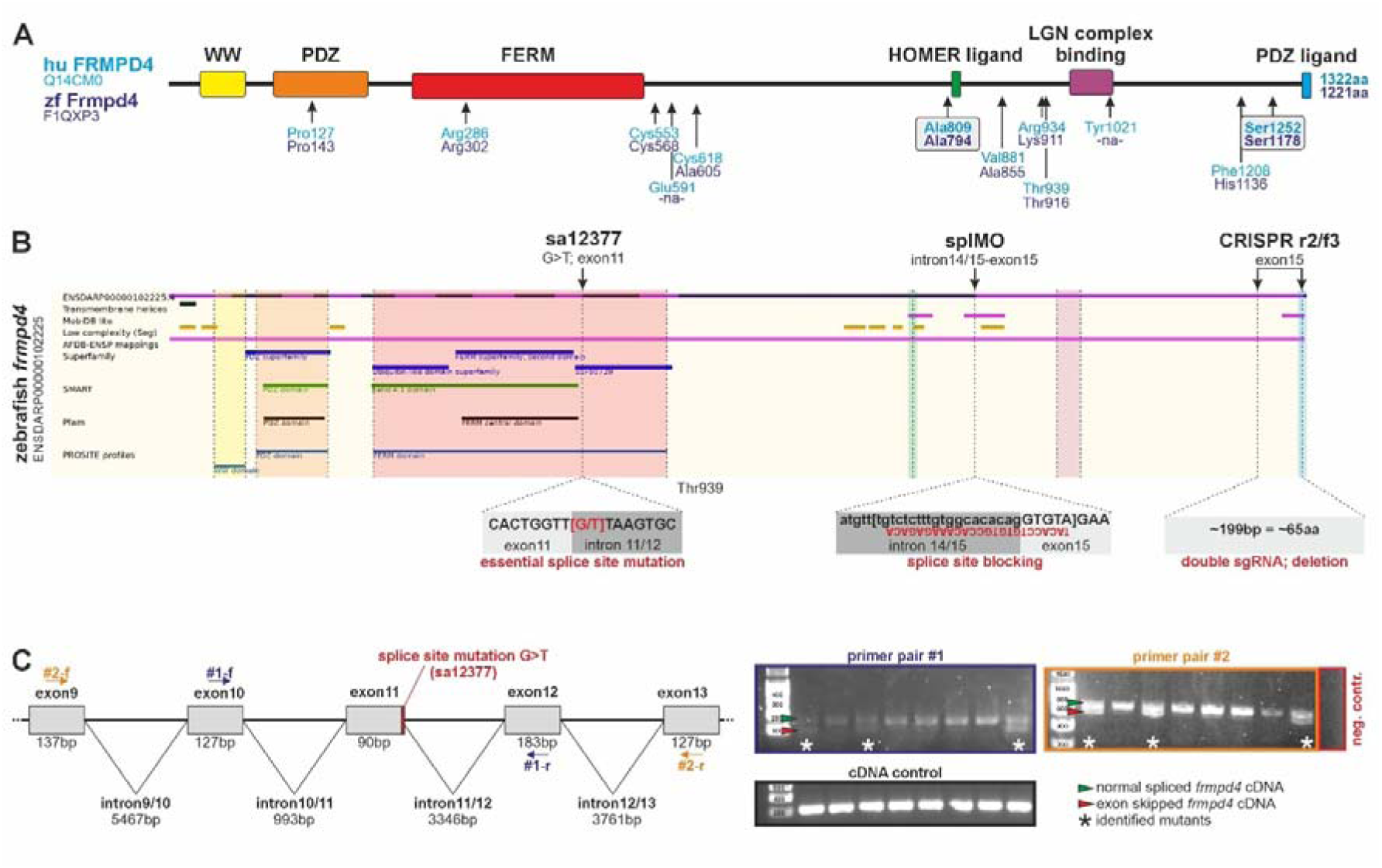
Summary of Frmpd4 domains in zebrafish. **(A)** Conservation of human variants in zebrafish. **(B)** Overview about used zebrafish knockdown reagents. **(C)** *frmpd4* locus overview and example of *frmpd4*^sa12377^ mutant identification by RT-PCR on cDNA from single embryos with two independent primer pairs spanning mutation site in exon11. Identified mutants (marked by asterisks) show an exon skipped PCR product (red arrowhead) in addition to a normal spliced product (green arrowhead). Amplification of *ef1a1* from the same cDNA was used as positive control. A PCR reaction without cDNA depicts a negative control.

**Supplementary Figure 8.**
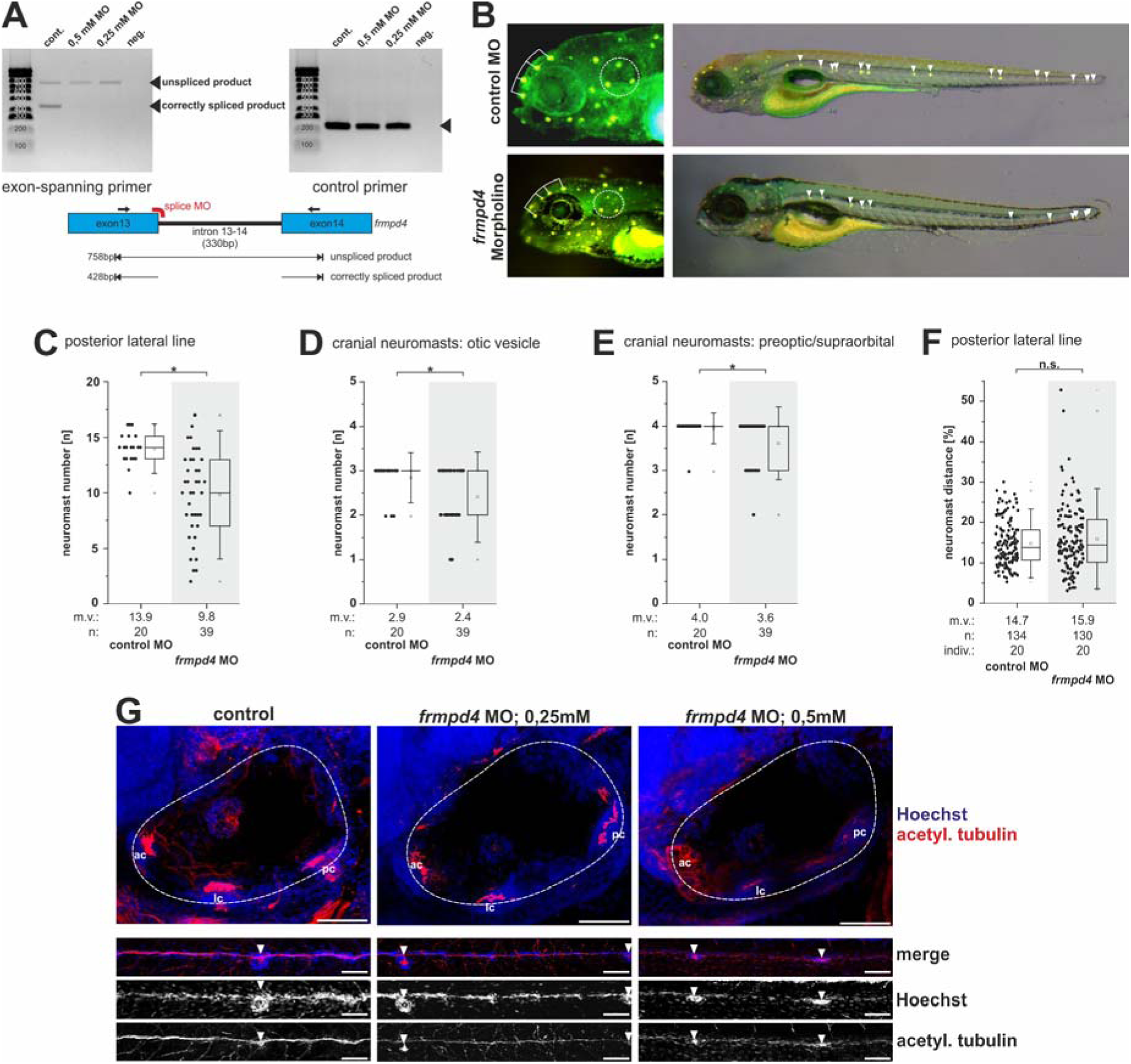
Loss of function of *frmpd4* in zebrafish results in neuromast alterations in the otic vesicle and in the posterior lateral line. **(A)** Functional test of *frmpd4* splice blocking Morpholino via RT-PCR (per condition 10 pooled embryos). Exon-spanning PCR primers indicate the loss of a correctly spliced *frmpd4* band after Morpholino injection, and gain of a larger, unspliced additional PCR product. PCR amplification of *ef1a1* were used as internal cDNA control. **(B)** DASPEI staining for neuromasts in *frmpd4* Morpholino injected knockdown embryos. Quantification of neuromast number in the posterior lateral line **(C)**, the otic vesicle **(D)** and the preoptical/supraorbital **(E)** indicated reduction after *frmpd4* knockdown (0.25mM Morpholino concentration, age: 4 dpf, comparison to embryos injected with standard MO). **(F)** Measurement of neuromast distance in the posterior lateral line indicated no significant change after *frmpd4* Morpholino knockdown. **(G)** Staining for acetyl tubulin after *frmpd4* Morpholino knockdown in 4 dpf larvae indicated similar to in *frmpd4^sa12377^* mutants loss of neuronal cells and axonal projections in ventral sensory patches of the otic vesicle and interference with correct neuromast formation in the posterior lateral line. Analyzed embryos: 2 wild type controls; 4 *frmpd4* MO 0.25mM; 2 *frmpd4* MO 0.5mM.

**Supplementary Figure 9.**
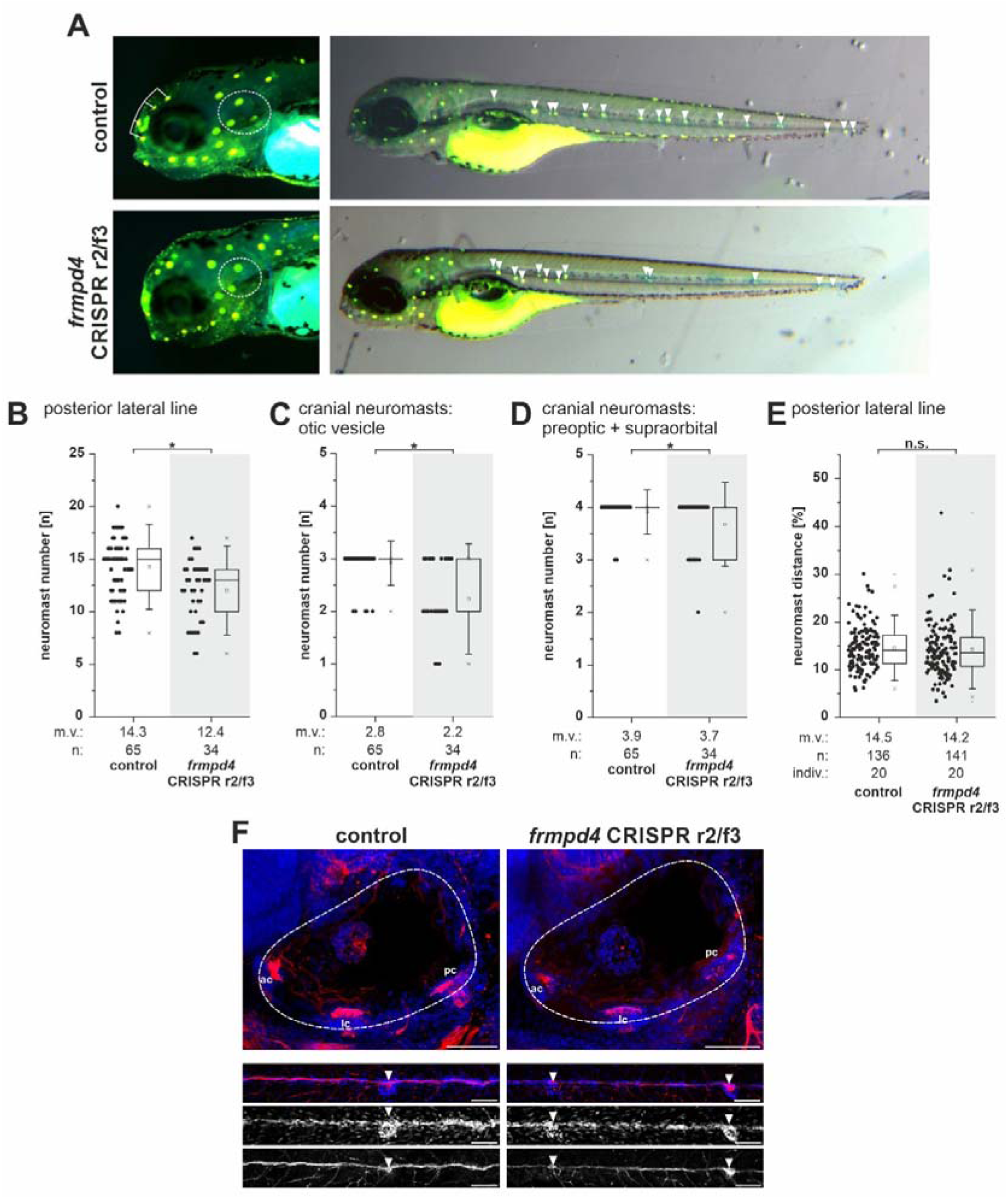
*frmpd4* CRISPant in zebrafish show only mild neuromast alterations in the otic vesicle and in the posterior lateral line. **(A)** DASPEI staining for neuromasts in CRISPants embryos (F0 generation, *frmpd4* sgRNA r2 and f3 injected). Quantification of neuromast number in the posterior lateral line **(B)**, the otic vesicle **(C)**, and the preoptical/supraorbital **(D)** indicated reduction in *frmpd4* CRISPants (4 dpf). **(E)** Measurement of neuromast distance in the posterior lateral line indicated no significant change after frmpd4 CRISPR or Morpholino knockdown. **(F)** Staining for acetylated tubulin in 4 dpf embryos indicated mild changes to neuronal cells and axonal projections in ventral sensory patches of the otic vesicle and not significant interference with correct neuromast spacing in the posterior lateral line after transient *frmpd4* CRISPR knockdown. Analyzed embryos: 2 wild type controls; 4 *frmpd4* CRISPants.

**Supplementary Figure 10.**
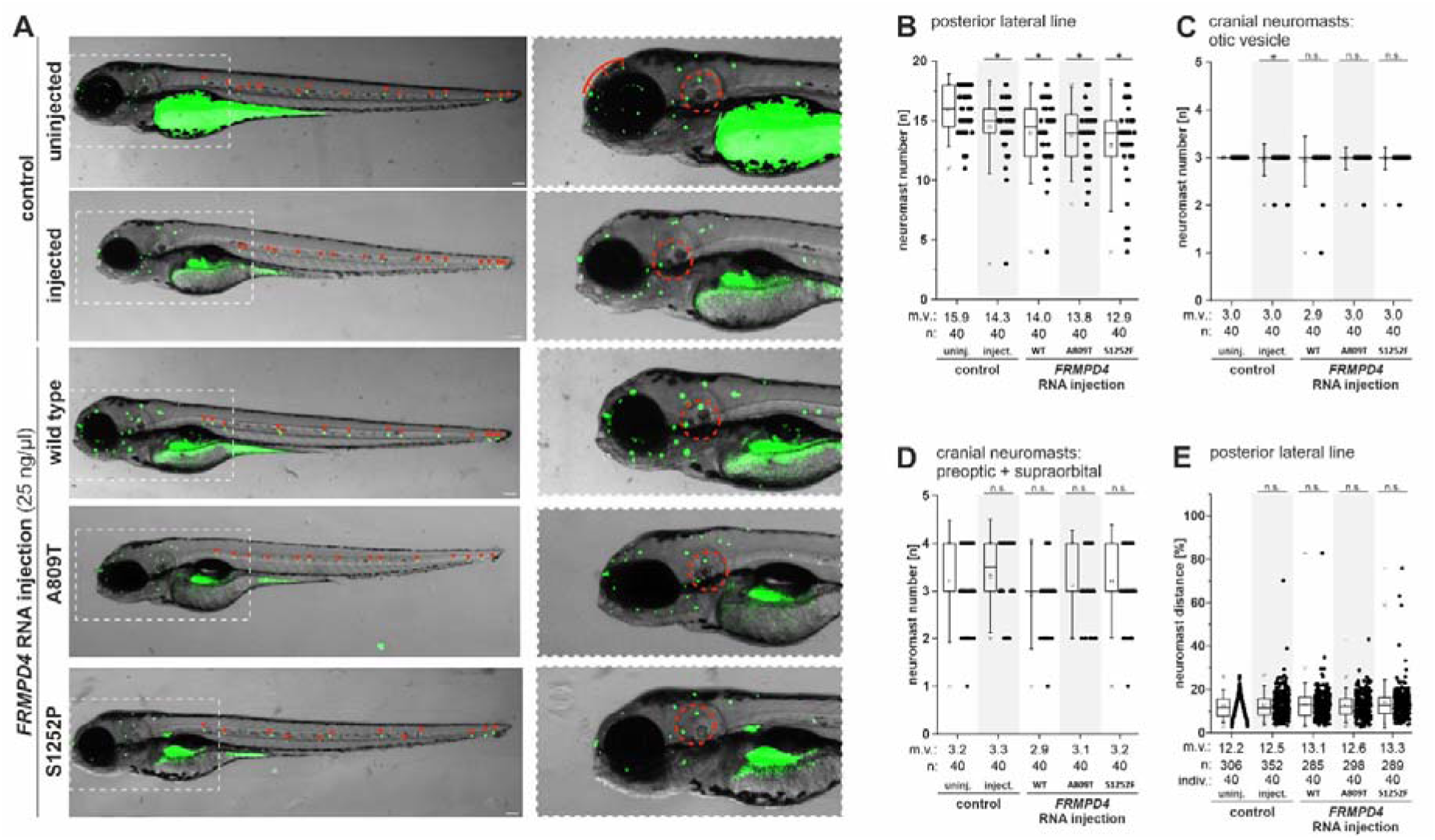
*FRMPD4* variant expression in zebrafish via RNA injection. **(A)** Brightfield and DASPEI overlay marking hair cells of 4 dpf zebrafish embryos injected with capped mRNAs coding for patient-specific *FRMPD4* variants. Quantification of neuromast number in the posterior lateral line **(B)**, the otic vesicle **(C)** and the preoptical/supraorbital **(D)**, and measurement of neuromast distance in the posterior lateral line **(E)** (Injection concentration: 25ng/ml RNA; injection controls: water instead of RNA). Regions of shown magnifications are outlined with white dashed lines. The otic vesicles are outlined with red dashed lines. DASPEI positive hair cells/ neuromasts of the pLL are marked with red arrowheads. m.v. – mean value; n – amount; indiv. – number of investigated individuals. Statistical significance as calculated by a two-tailed Mann-Whitney U test in comparison to uninjected controls. Statistical significance is depicted as * for U ≤ 0.05, n.s. mark not significant different groups. Scale bars indicate 100µm.

### Supplementary Movies

**Supplementary Movie 1:** Examples of a startle response reaction to given sound stimulus, one individual per *frmpd4^sa12377^* genotype, low dB sound stimulus (Frmpd4 lowbD.mp4)

**Supplementary Movie 2:** Examples of a startle response reaction to given sound stimulus, one individual per *frmpd4^sa12377^* genotype, high dB sound stimulus (Frmpd4 highbD.mp4)

